# Artificial Intelligence Methods to Detect Heart Failure with Preserved Ejection Fraction (AIM-HFpEF) within Electronic Health Records: An equitable disease prediction model

**DOI:** 10.1101/2024.12.24.24319603

**Authors:** Jack Wu, Dhruva Biswas, Samuel Brown, Matthew Ryan, Brett Bernstein, Brian Tam To, Tom Searle, Maleeha Rizvi, Natalie Fairhurst, George Kaye, Ranu Baral, Dhanushan Vijayakumar, Daksh Mehta, Narbeh Melikian, Daniel Sado, Gerald Carr-White, Phil Chowienczyk, James Teo, Richard JB Dobson, Daniel I Bromage, Thomas F Lüscher, Ali Vazir, Theresa A McDonagh, Jessica Webb, Ajay M Shah, Kevin O’Gallagher

## Abstract

**Background and aims:** Heart Failure with Preserved Ejection Fraction (HFpEF) accounts for approximately half of all heart failure cases, with high levels of morbidity and mortality. However, most cases of HFpEF are undiagnosed as conventional risk scores underestimate risk in non-White populations. Our aim was to develop and validate a diagnostic prediction model to detect undiagnosed HFpEF, AIM-HFpEF.

**Methods:** We applied natural language processing (NLP) and machine learning methods to routinely collected electronic health record (EHR) data from a tertiary centre hospital trust in London, UK, to derive the AIM-HFpEF model. We then externally validated the model and performed benchmarking against existing HFpEF prediction models (H2FPEF and HFpEF-ABA) for diagnostic power in patients of non-white ethnicity and patients from areas of increased socioeconomic deprivation.

**Results:** An XGBoost model combining demographic, clinical and echocardiogram data showed strong diagnostic performance in the derivation dataset (n=3170, AUC=0.88, [95% CI, 0.86-0.91]) and validation cohort (n=5383, AUC: 0.88 [95% CI, 0.87-0.89]). Diagnostic performance was maintained in patients of non-White ethnicity (AUC=0.88 [95% CI, 0.84-0.93]) and patients from areas of high socioeconomic deprivation (AUC=0.89 [95% CI, 0.84-0.94]). and AIM-HFpEF performed favourably in comparison to H2FPEF and HFpEF-ABA models. AIM-HFpEF model probabilities were associated with an increased risk of death, hospitalisation and stroke in the external validation cohort (P<0.001, P=0.01, P<0.001 respectively for highest versus middle tertile).

**Conclusion:** AIM-HFpEF represents a validated equitable diagnostic model for HFpEF, which can be embedded within an EHR to allow for fully automated HFpEF detection.

## Introduction

Heart Failure with Preserved Ejection Fraction (HFpEF) accounts for approximately half of all heart failure (HF) cases and is associated with significant healthcare costs, morbidity, and mortality. Early diagnosis is important and has prognostic benefit ^1^. However, despite the high prevalence of HFpEF, most cases remain undiagnosed ^2^ and therefore a high proportion of patients do not benefit from specialist cardiology care, evidence-based therapies, and eventually improved outcomes. ^3,4^

Strategies have been developed to improve HFpEF detection, including the H2FPEF scoring system ^5^. The H2FPEF score is validated for the diagnosis of HFpEF, but requires an *a priori* suspicion of HFpEF. More recently, the HFpEF-ABA score, based on clinical features alone with no cardiac imaging features, has been developed as a screening tool to identify possible HFpEF cases and guide the need for specialist cardiac imaging and clinical evaluation ^6^.

HFpEF is disproportionately under-diagnosed particularly in patients of non-white race and ethnicity ^7^. Indeed, Black and Asian patients with HFpEF have different patterns of comorbidity to White patients, including features used in current HFpEF diagnostic systems such as atrial fibrillation and body mass index.

The widespread deployment of electronic health record (EHR) platforms provides the potential to enable access to a wide range of routinely collected clinical data in a fraction of the time taken to perform manual case record completion. EHR-based diagnostic approaches lend themselves to automation, removing the need for clinician-initiated suspicion of disease and therefore potentially decreasing the risk of bias. Moreover, advances in artificial intelligence (AI) methods allow for the capture of both structured and unstructured data, including AI-based detection of clinical concepts from free text via natural language processing (NLP) ^8–11^ with potentially less data missingness in populations less engaged with health services. We have previously used these methods to detect undiagnosed HFpEF from the EHR, finding that less than 10% of all cases of HFpEF have a clinician-assigned diagnosis, while the remaining 90% are undiagnosed. ^2^

The aim of this study was to develop and externally validate a prediction model for detecting HFpEF by applying artificial intelligence methods to routinely collected data from the EHR of two independent, ethnically diverse cohorts of HFpEF patients, including a representative distribution of cases of Confirmed and Undiagnosed HFpEF. A further aim was to assess the performance of the prediction model across racial and ethnicity groups and in patients from socially deprived areas.

## Methods

### Ethical Considerations

This project operated under London South-East Research Ethics Committee approval (18/LO/2048) granted to the King’s Electronic Records Research Interface (KERRI) and London Dulwich Research Ethics Committee approval (19/LO/1957), which did not require written informed patient consent. The study complies with the Declaration of Helsinki. Patients were consulted on the study via a dedicated patient and public involvement (PPI) meeting held during the study design phase. A formal protocol was not published and the study was not registered.

### Participating Centres

King’s College Hospital NHS Foundation Trust (KCH) and Guy’s and St Thomas’ NHS Foundation Trust (GSTT) are two large, multi-site tertiary hospitals in London, UK, providing specialist cardiology services and a dedicated heart failure service open to referral by any physician.

### Study Design and inclusion criteria

At each participating centre, we established a registry comprising a retrospective anonymised database of adult patients with a clinical diagnosis of HF documented within the EHR between 2010-2022. **Figure 1** shows the schematic diagram of the study. Patients were included if they had two or more mentions of a “heart failure” (HF) diagnosis in the clinical text as determined by a well-validated NLP pipeline ^12–15^. Specifically, a random sample of 100 HFpEF patients identified by the NLP pipeline were manually validated for this study and 100% of them were true positive. Patients were included regardless of nature of clinical episode (inpatient or outpatient). Both structured and unstructured portions of the echocardiogram report were used to extract LVEF data of patients and other relevant echocardiographic parameters. Patients with a clinical diagnosis of HF and LVEF ≥50% were categorised into one of the following 2 groups:

1. Confirmed HFpEF: Clinician-assigned diagnosis of HFpEF
2. Undiagnosed HFpEF (meeting the ESC diagnostic criteria): Patients with HF, LVEF≥50%, and imaging/biochemical evidence of diastolic dysfunction meeting the ESC diagnostic criteria ^16^ but who have not received a HFpEF diagnosis.

**Figure 1.**
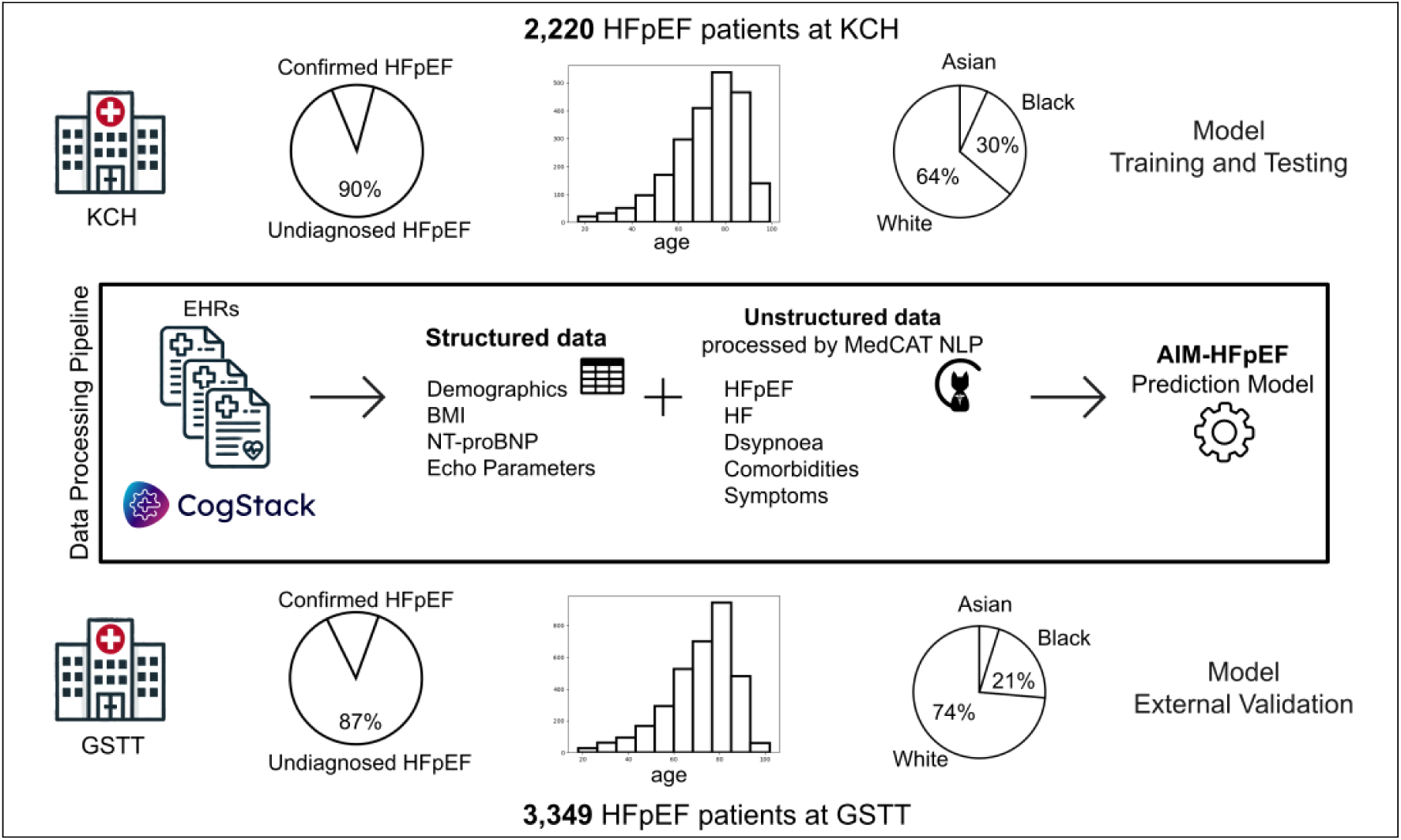
Schematic diagram of the study.

Patients were excluded if at any point they had an echocardiogram with LVEF <50%. Patients with a clinical diagnosis of HF and LVEF≥50% on echocardiography, but not meeting ESC diagnostic criteria and who have not received a HFpEF diagnosis were excluded, as were patients with an alternative diagnosis (severe valvular heart disease, hypertrophic cardiomyopathy, restrictive cardiomyopathy, constrictive pericarditis, and cardiac amyloidosis). **Supplementary Table 1** contains the SNOMED terms used in the classification of included and excluded patients. Patients needed to have the echocardiogram report within one year of clinical diagnosis of HF, otherwise they were excluded.

At each site we also established a registry of non-cardiac dyspnoea patients to act as control groups. Patients in the control group needed (1) two or more mentions of dyspnoea in the clinical text as determined by NLP; (2) an echocardiogram report (LVEF≥50%) within one year of the dyspnoea mentions; and (3) no mentions of HF diagnosis in the clinical text. Patients with severe valvular heart disease, hypertrophic cardiomyopathy, restrictive cardiomyopathy, constrictive pericarditis, or cardiac amyloidosis were also excluded from the control group.

### Prediction Model Development

The dataset collected from KCH was used as the derivation cohort, with 80% of the data used for training and 20% for testing, while the GSTT dataset was used for external validation. All model development and evaluation were performed using Python version 3.10.12.

We derived prediction models using four different methods: Logistic Regression (LR), Support Vector Machine (SVM), Random Forest (RF) and XGBoost (XGB). The machine learning models were built and evaluated using the ‘scikit-learn’ (for LR, SVM and RF) and ‘xgboost’ (for XGB) python packages. Hyperparameter tuning for each model was performed through grid search and 10-fold cross-validation using the training dataset only. The ‘StandardScaler’ from ‘scikit-learn.preprocessing’ was applied to the data before model training for consistent scaling across echocardiograph parameter features for LR, SVM and RF models. Mean imputation was applied to the missing values for the three (LR, SVM and RF) machine learning models, missForest imputation was also tested and the correlation of the predicted probabilities from both methods were greater than 98%. XGB can handle scaling and missing values internally. Model evaluation was based on metrics including accuracy, precision, recall, F1 score, and AUC-ROC, calculated using functions from ‘scikit-learn.metrics’. The comparison of AUCs was performed using a python implementation of the DeLong test ^17^. We selected the model which had the best overall performance for the Results section while the performances for non-selected models are reported in **Supplementary Table 2**.

### Feature Engineering

We aimed to produce a model based on the parameters included in the ESC criteria for diagnosing HFpEF. Features representing key demographic, echocardiographic, comorbid, and symptomatic factors commonly associated with HFpEF are used to construct the model and they are extracted from both structured and unstructured data. Features from structured data include age, sex, BMI, NTproBNP and echocardiographic parameters (E/e’, LA volume, LA volume indexed, IVSd, LVPWD, LVEDD, LV V1 max, LV V1 max PG, LV mass, LV mass indexed, and PASP), while comorbidities (diabetes mellitus type 1, diabetes mellitus type 2, ischemic heart disease, myocardial infarction, cerebrovascular accident, hypertensive disorder, transient ischemic attack, atrial fibrillation, pulmonary hypertension, kidney disease, and angina) and symptoms (dyspnoea at rest, chest pain, dizziness, and syncope) were extracted from unstructured clinical notes through natural language processing (NLP) using MedCAT ^13^ within the CogStack platform ^18^. Echocardiographic parameters included in the ESC criteria were prioritized in the model, while other parameters with more than 30% missingness were excluded. Comorbidities and symptoms are represented as binary features, with a positive value indicating the presence of the comorbidity or symptom before the first mention of HF (for the HFpEF group) or dyspnoea (for the control group) in the EHR while a negative value indicates the absence of the condition. Dyspnoea was excluded as a feature in the model since it was part of the inclusion criteria for control patients, resulting in 100% of the control group presenting with dyspnoea. The full model includes 30 features in total and can be executed automatically on the CogStack platform using routinely collected EHR data.

A generalised linear model (GLM) from the ‘statsmodels’ package was used to simplify the full prediction model through feature selection using the KCH training dataset, retaining the top 10 features ranked by significance based on their p-values. These selected features were then used to construct a simplified prediction model. In the simplified model, diabetes mellitus type 1 and type 2 were combined into a single feature. The simplified model can be useful for manually inputting feature values in the form of an online application.

### Model explainability

A Shapley Additive Explanations (SHAP) graph was plotted for the full model using the ‘shap’ python library (version 0.45.1) ^19^ to show the importance and values of each feature contributing to the prediction outcome.

### Comparison with H2FPEF and HFpEF-ABA

For comparison with the H2FPEF score, we followed the more precise version of the H2FPEF probability using the formula provided in Reddy et al. (2018) ^5^, specifically the online calculator from the supplemental material. The formula requires five key variables: BMI, atrial fibrillation (AF), pulmonary artery systolic pressure (PASP), age, and filling pressure (E/e’). We also compared the results with the point-based version of the H2FPEF score (which requires six variables with antihypertensive drugs added) in **Supplementary Table 2**. Patients with any missing values for these key variables were excluded from the comparison to ensure accurate probability estimation.

For comparison with the HFpEF-ABA score, we used the formula provided in Reddy et al. (2024) ^6^, specifically from “Extended Data Table 4 Regression equations for clinical variable models”. This model requires three variables: age, BMI, and atrial fibrillation (AF). Similar to the comparison with H2FPEF score, patients with missing values for any of these three variables were excluded from the comparison.

### Subgroup analysis

In the subgroup analysis, we examined two patient subgroups: (1) non-White individuals based on self-ascribed ethnicity ^2^ and (2) those with low socioeconomic status, as assessed by the English Indices of Multiple Deprivation 2019 (IMD). The IMD was determined using postcodes of the patients. Patients were classified as having low IMD if their postcodes fell within the most deprived quintiles according to the national index. The model’s performance was evaluated separately in the two subgroups to assess potential variations in predictive accuracy for HFpEF based on ethnicity and socioeconomic status.

### Prediction Model Output and Calibration

The machine learning models output the probability of HFpEF for each patient. For consistency, the cut-off value for a positive HFpEF prediction is set at 0.5 when computing the accuracy, precision and recall values in **Supplementary Table 2**. The same threshold is used when comparing those metrics with the H2FPEF and HFpEF-ABA scores. In practice, the threshold can be adjusted according to individual treatment goals and preferences ^6^. Calibration of the prediction probabilities were assessed graphically using calibration curves produced by the ‘calibration_curve’ function in ‘scikit-learn’ package.

### Outcome data

Mortality data were obtained from death notification letters in the EHR system and the master demographic patient indices of the hospitals (synchronised with NHS Spine for demographics). Hospitalisations were estimated by the number of discharge notifications in the EHR in the study timeframe i.e. 2010-2022. Diagnoses of myocardial infarction and stroke were recorded as outcome data if they occurred after the first mention of HF (for the HFpEF group) or dyspnoea (for the control group) in the EHR.

## Results

We identified 2,220 HFpEF patients (231 [10%] confirmed HFpEF and 1,989 [90%] undiagnosed HFpEF) at KCH and 3,349 HFpEF patients (430 [13%] confirmed HFpEF and 2,919 [87%] undiagnosed HFpEF) at GSTT. There were 950 and 2,034 non-HF patients with dyspnoea at KCH and GSTT, respectively, for the control groups. **Table 1** shows the baseline characteristics of the patients.

**Table 1.**
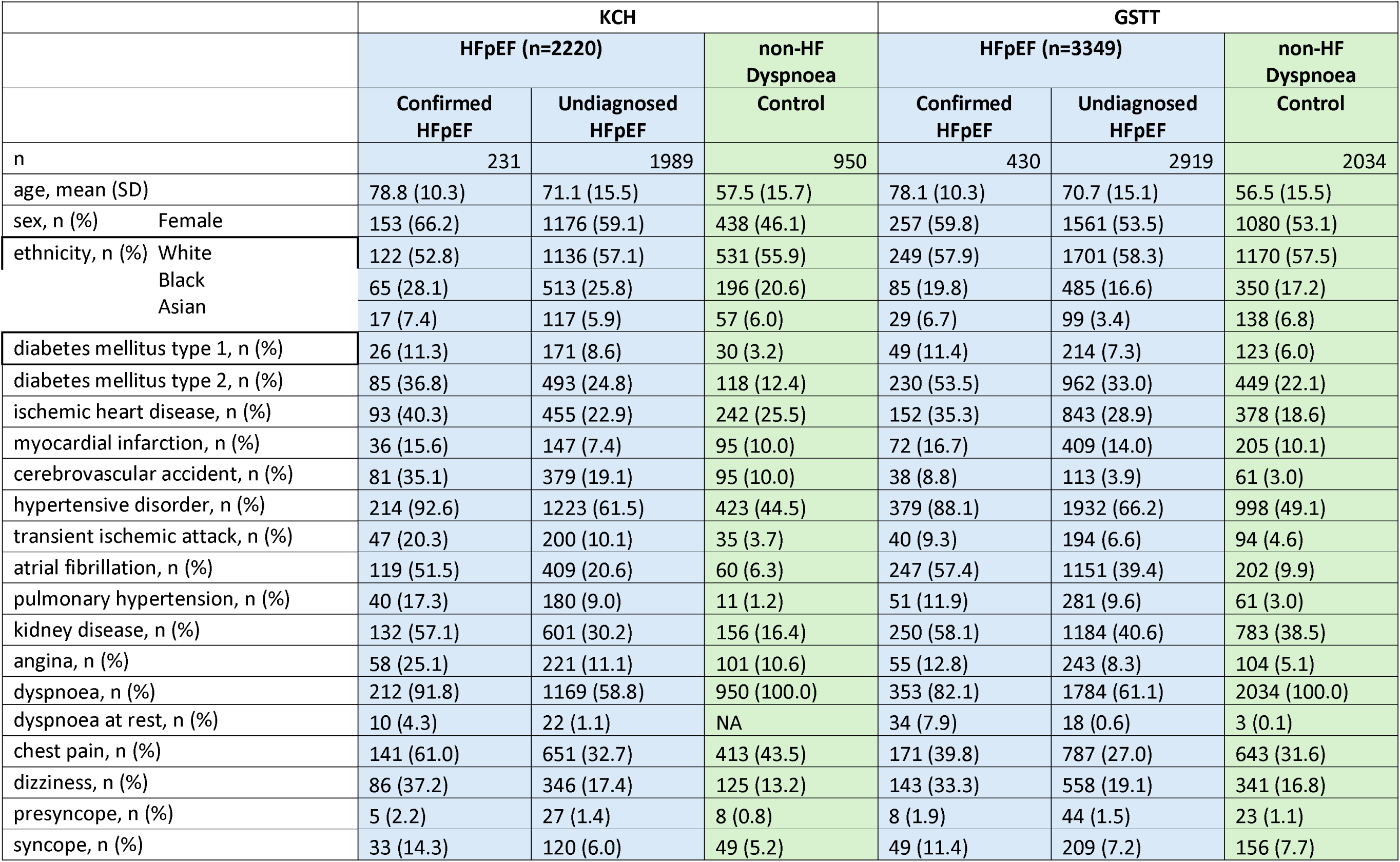
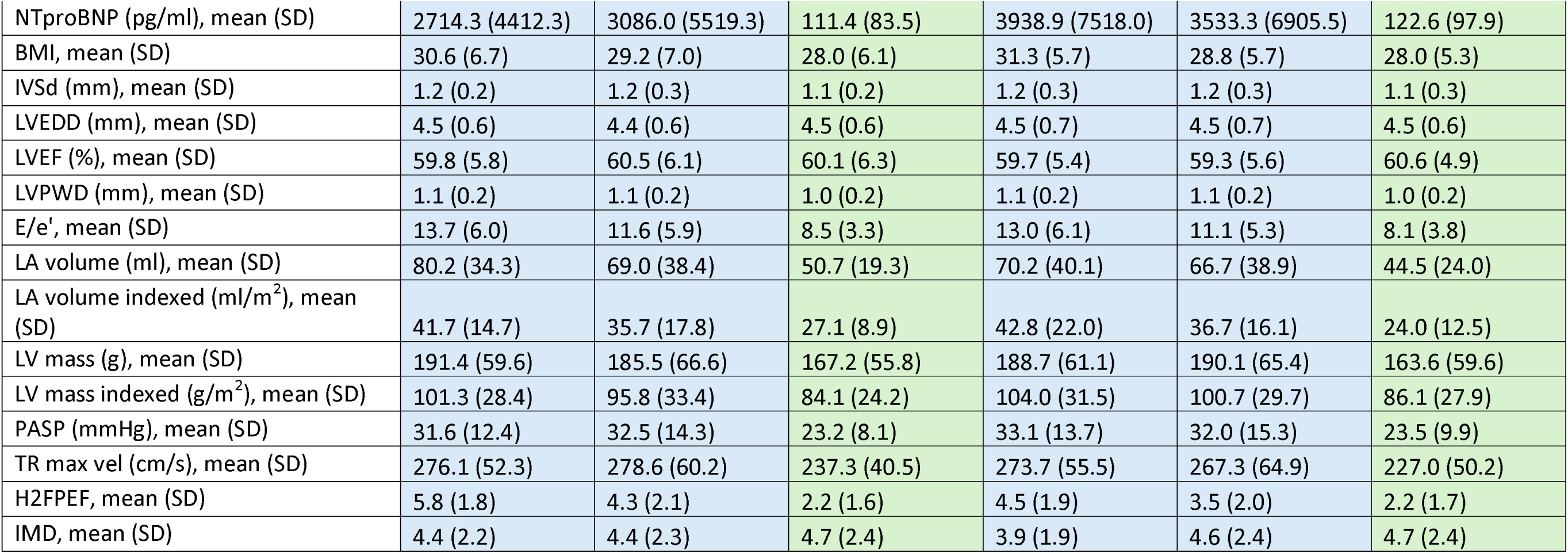
Baseline characteristics (demographics, comorbidities, symptoms, echocardiograph parameters) of patients.

We randomly divided the KCH patients into 80% for training (a total of 2,536 patients: 177 confirmed HFpEF, 1,599 undiagnosed HFpEF, and 760 control) and 20% for testing (a total of 634 patients: 54 confirmed HFpEF, 390 undiagnosed HFpEF, and 190 control), stratified by HFpEF and non-HF groups. The GSTT patients were used for external validation.

Among the four machine learning models: logistic regression (LR), random forest (RF), support vector machine (SVM), and XGBoost (XGB), the XGB model achieved the highest performance, with an AUC-ROC of 0.8910 (95% CI, 0.8665-0.9154) in the KCH testing cohort, outperforming the other models (LR:0.8204 [95% CI, 0.7868-0.8540], RF: 0.8472 [95% CI, 0.8161-0.8784], SVM:0.8386 [95% CI, 0.8063-0.8709]). Therefore, we focused on the XGBoost model for further analysis and validation. The full report of the performances of all the models is shown in **Supplementary Table 2**.

We employed SHAP analysis to understand the contribution of individual features to the model’s predictions from XGB as shown in **Figure 2 (a)**. The top contributing features include NTproBNP, age, PASP, LA volume and BMI. Age, PASP and BMI are also included in the H2FPEF formula but the XGB model has no prior information of this.

**Figure 2.**
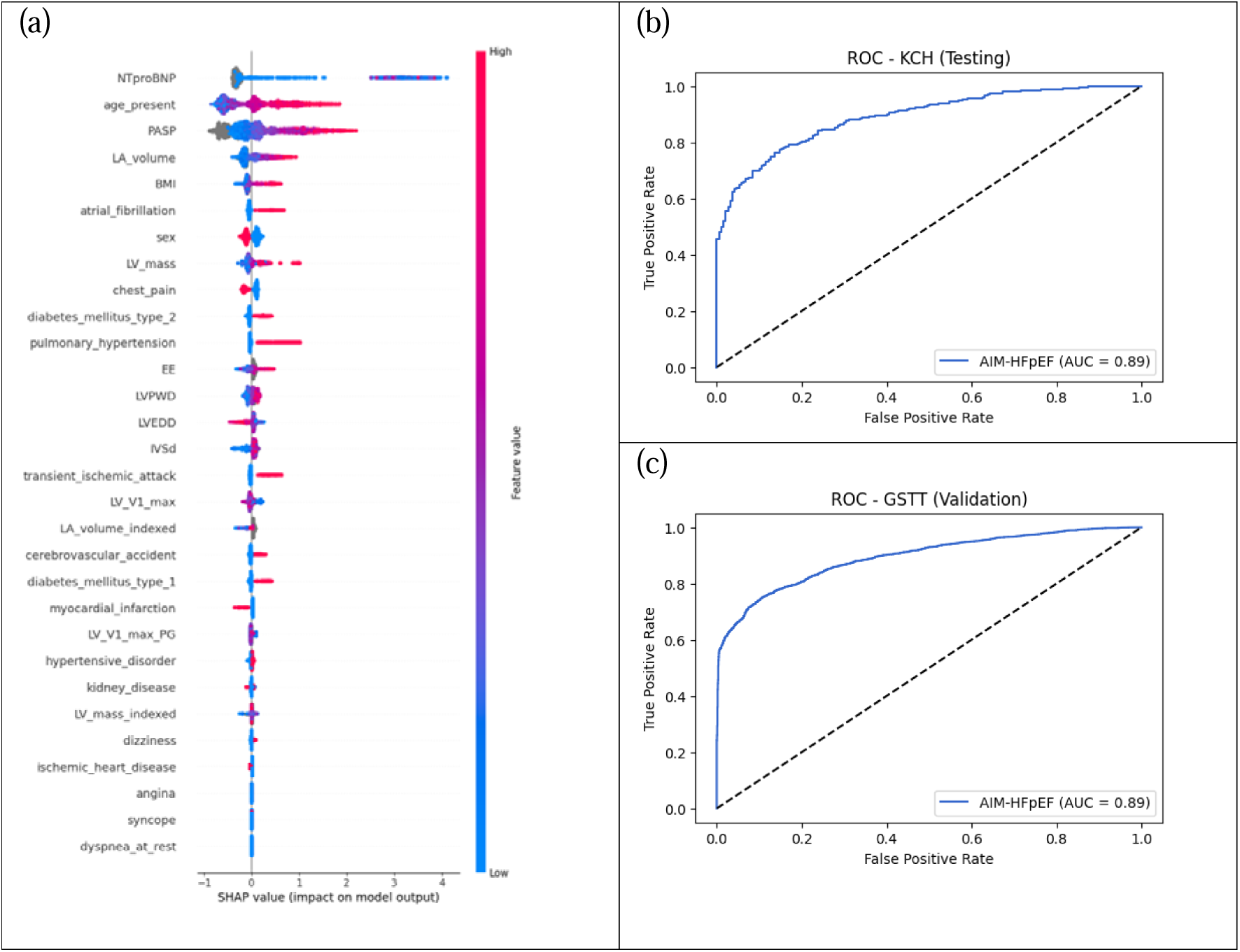
(a) SHAP graph for the XGB AIM-HFpEF full model, the y-axis is the list of features ranked by their importance with the most important feature at the top. The x-axis shows the SHAP values which indicate the impact of each feature on the model’s prediction. Positive SHAP values show to a tendency of positive HFpEF prediction. The colour reflects the values of the features (for continuous features: red represents higher values and blue lower values; for sex: red represents male and blue female; for binary features like comorbidities and symptoms: red represents positive and blue negative). (b) ROC curve of KCH testing cohort. (c) ROC curve of GSTT validation cohort.

**Figure 2(b) and (c)** shows the Receiver Operating Characteristic (ROC) curves for the XGB prediction model on the KCH testing dataset and GSTT validation dataset, respectively. The AUC for the GSTT validation dataset was 0.8934 (95% CI, 0.8852-0.9016) for the XGB model.

### Simplified Model

While the SHAP graph can help us to understand how the XGB model predicts the patients, in order to identify features with the strongest statistical relationship with the prediction outcome, a Generalized Linear Model (GLM) analysis was performed to select the top 10 features for a simplified model. **Table 2** shows the results of the GLM with the top selected features and their coefficients and p-values associated with HFpEF prediction using the KCH training dataset. All the top 10 features have a p-value of less than 0.02 indicating that they are statistically significant predictors.

**Table 2.** GLM selected features.

| Table 2. GLM selected features. |  |  |
| --- | --- | --- |
| Feature | Coefficient | P-Value |
| Age at presentation | 0.5640 | <0.0001 |
| PASP | 0.5713 | <0.0001 |
| Sex | -0.2801 | <0.0001 |
| NT-proBNP | 1.0217 | <0.0001 |
| Atrial fibrillation | 0.2816 | 0.0002 |
| LVEDD | -0.3566 | 0.0024 |
| Transient ischemic attack | 0.2012 | 0.0033 |
| Diabetes mellitus | 0.1804 | 0.0046 |
| BMI | 0.1790 | 0.0113 |
| LA volume | 0.4207 | 0.0139 |

A simplified version of the machine learning models was next built using the 10 features identified by GLM. The XGB model again performed the best (AUC-ROC of 0.8845 [95% CI, 0.8592-0.9097]) among the four models (LR: 0.8025 [95% CI, 0.7678-0.8373], RF: 0.8379 [95% CI, 0.8051-0.8707], SVM: 0.8205 [95% CI, 0.7852-0.8559]) for the KCH testing cohort. The AUC of the simplified model is slightly less than that in the full model (AUC of 0.8809 [95% CI, 0.8721-0.8897] for the GSTT validation cohort), but it is more usable in practice. The individual performances of the model on the confirmed HFpEF and undiagnosed HFpEF groups are shown in **Supplementary Figure 1**. The calibration curves for the two HFpEF groups in the GSTT validation cohort is shown in **Supplementary Figure 2**. The simplified XGB model is used for subsequent comparison and subgroup analysis.

### Comparison with H2FPEF and HFpEF-ABA scores

To evaluate the performance of the simplified XGB model in comparison with established clinical scoring systems, we compared it to the widely used H2FPEF score and the more recently published HFpEF-ABA score. In the KCH testing cohort (n=634), 182 (29%) patients had complete data for all five variables (age, BMI, PASP, E/e’, and AF) required to calculate the H2FPEF probability (AUC, AIM-HFpEF: 0.8751 [95% CI, 0.8225-0.9276], H2FPEF: 0.7873 [95% CI, 0.7075-0.8672], p=0.0064), while 483 (76%) patients had all three variables (age, BMI, and AF) necessary for the HFpEF-ABA score calculation (AUC, AIM-HFpEF: 0.8792 [95% CI, 0.8493-0.9091], HFpEF-ABA: 0.7425 [95% CI, 0.6949-0.7901], p<0.0001). In the GSTT validation cohort (n=5,621), 967 (17%) patients had the full set of variables to compute the H2FPEF probability (AUC, AIM-HFpEF: 0.8746 [95% CI, 0.8510-0.8982]), H2FPEF: 0.7805 [95% CI, 0.7505-0.8105], p<0.0001) and 3,735 (66%) patients had the variables required for the HFpEF-ABA score (AUC, AIM-HFpEF: 0.8788 [95% CI, 0.8665-0.8912], HFpEF-ABA: 0.7624 [95% CI, 0.7462-0.7787], p<0.0001). **Figure 3 (a)** presents the ROC curves comparing the performance of the simplified XGB model, H2FPEF score, and HFpEF-ABA score in the GSTT validation cohort. Comparison for the KCH testing cohort is shown in **Supplementary Figure 3(a)**.

**Figure 3.**
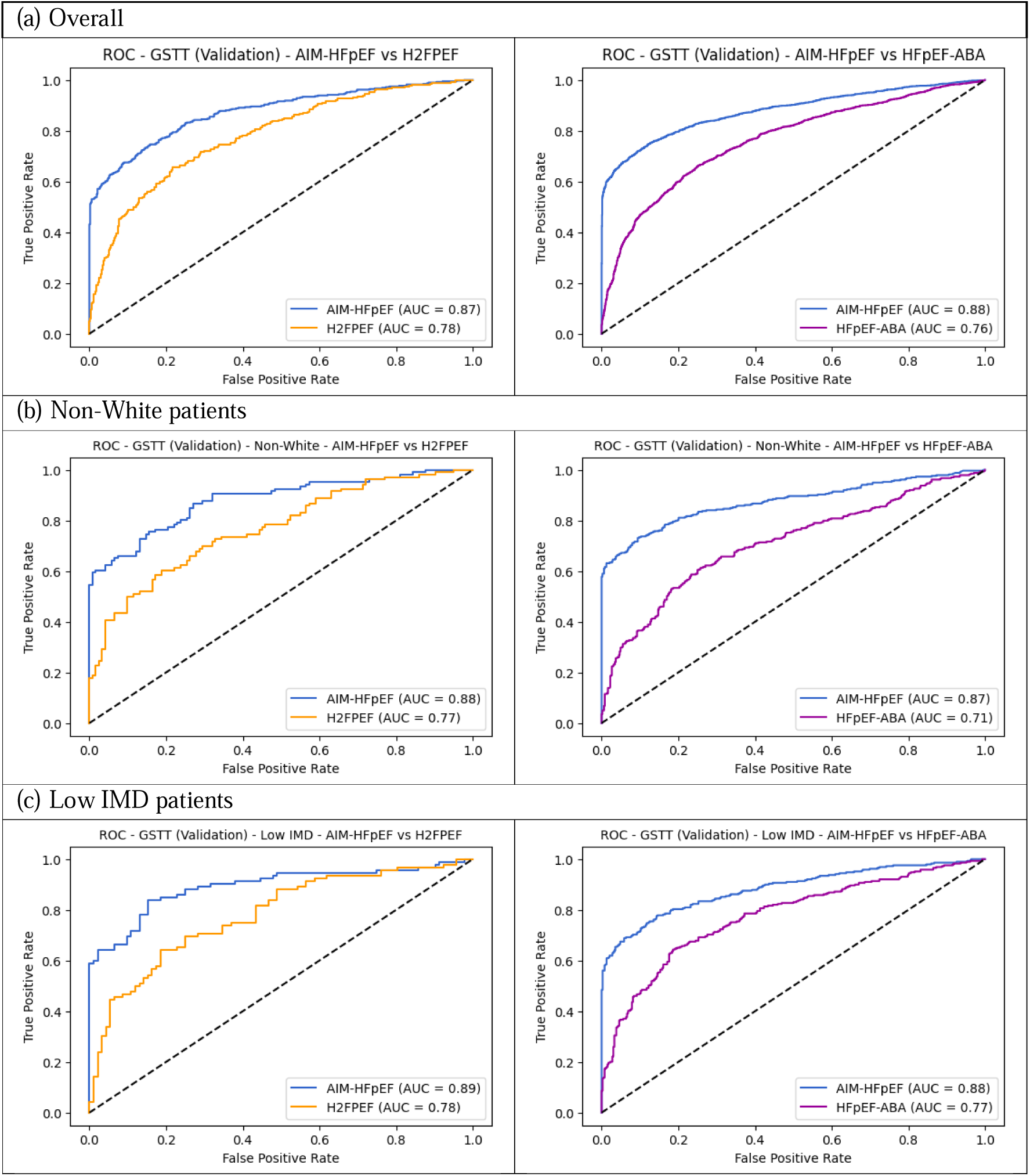
Comparisons of Simplified XGB, H2FPEF and HFpEF-ABA in the GSTT validation cohort.

### Subgroup analysis

In GSTT validation cohort (n=5,621), there were 1,282 (23%) non-White patients and 1,136 (20%) patients from the lowest quintile of IMD.

Non-White patients: In the GSTT validation cohort, the number of patients included in the comparison for the H2FPEF and HFpEF-ABA scores were 249 (AUC, AIM-HFpEF: 0.8812 [95% CI, 0.8365-0.9259], H2FPEF: 0.7681 [95% CI, 0.7071-0.8291], p<0.0001) and 923 (AUC, AIM-HFpEF: 0.8744 [95% CI, 0.8477-0.9012], HFpEF-ABA: 0.7101 [95% CI, 0.6734-0.7468], p<0.0001), respectively. **Figure 3 (b)** shows the ROC curves for the GSTT validation cohort. Comparisons for the KCH testing cohort is shown in **Supplementary Figure 3(b)**.

Low IMD patients: In the GSTT validation cohort, the corresponding numbers were 195 (AUC, AIM-HFpEF: 0.8914 [95% CI, 0.8423-0.9406], H2FPEF: 0.7793 [95% CI, 0.7127-0.8459], p=0.0003) and 726 (AUC, AIM-HFpEF: 0.8829 [95% CI, 0.8557-0.9100], HFpEF-ABA: 0.7745 [95% CI, 0.7383-0.8107], p<0.0001), respectively. **Figure 3 (c)** shows the ROC curves for the GSTT validation cohort. Comparisons for the KCH testing cohort is shown in **Supplementary Figure 3(c)**.

#### Outcome analysis

In the outcome analysis, we selected patients with a predicted probability ≥90% of having HFpEF from the models and investigated their all-cause mortality over a 5-year period. In the KCH testing cohort, the AIM-HFpEF model identified 223 patients and 89 (40%) of them died within 5 years of HF diagnosis. H2FPEF identified 32 patients and 13 (33%) died within 5 years, while HFpEF-ABA identified 87 and 29 (33%) of them died. In the GSTT validation cohort, the numbers are AIM-HFpEF: 2272 and 957 (42%); H2FPEF: 105 and 53 (50%); HFpEF-ABA: 606 and 193 (32%). Kaplan-Meier (KM) curves of the outcome analysis for all-cause mortality, MI and stroke are shown in **Supplementary Figure 4**. We also investigated whether the AIM-HFpEF model can predict outcome by dividing the GSTT validation cohort into three groups based on AIM-HFpEF predicted probability tertiles in Figure 4. In the overall cohort, AIM-HFpEF produced probabilities that were associated with an increased risk of death (p<0.0001), stroke (p=0.01) and hospitalisation (p<0.0001) when comparing the highest tertile to the middle tertile. In the undiagnosed cohort, AIM-HFpEF probabilities were associated with an increased risk of death (p<0.0001) and hospitalisation (p<0.0001).

**Figure 4.**
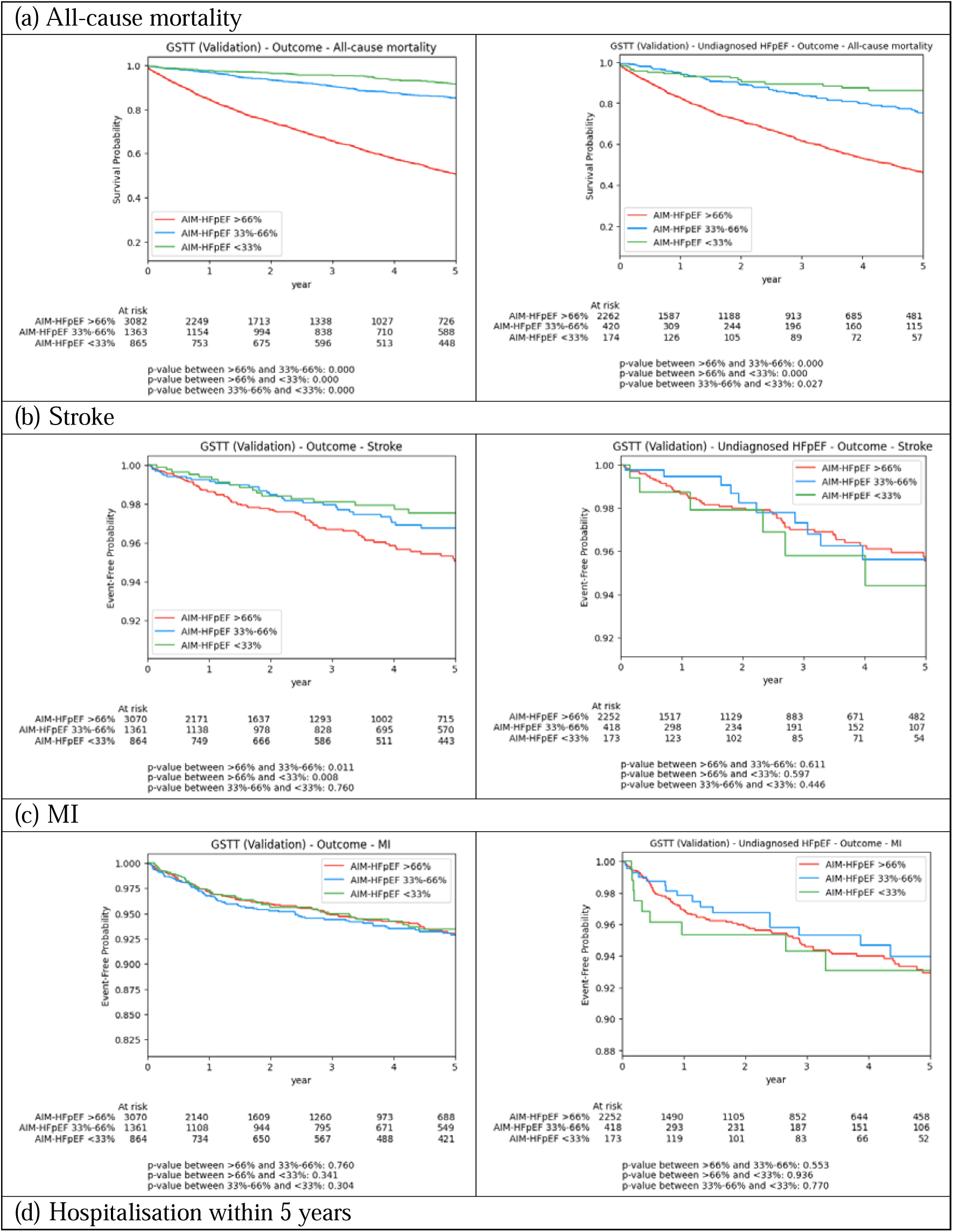
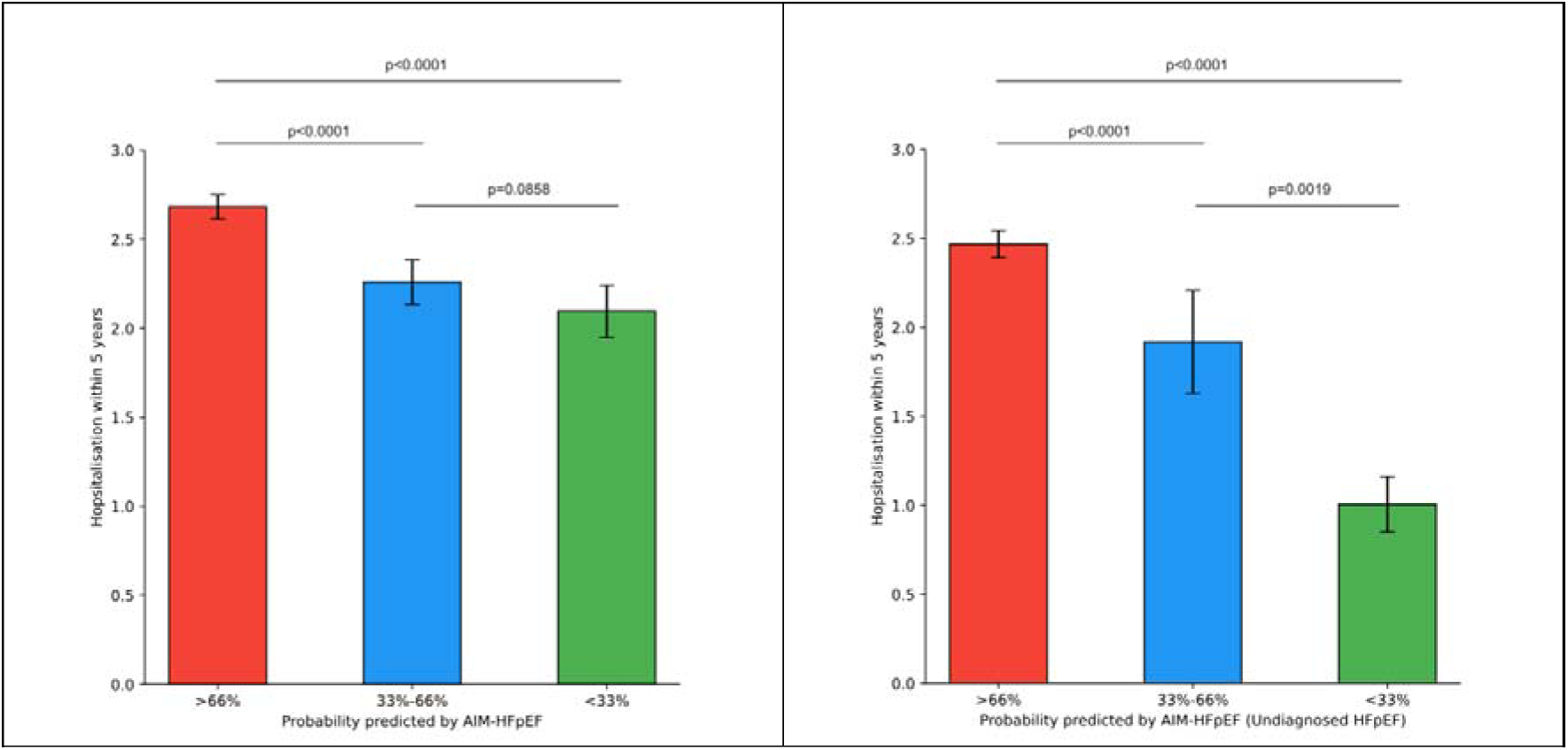
Outcome of GSTT validation cohort divided into three groups based on AIM-HFpEF predicted probability tertiles (left: overall cohort, right: undiagnosed HFpEF).

## Discussion

Using well-validated NLP and machine learning techniques applied to routinely collected data from the EHR, we have developed the AIM-HFpEF predictive model to accurately detect patients with HFpEF. Importantly from a health equity point of view and to address algorithmic bias ^20^, it performs well in patients of non-White ethnicity and in patients from areas of higher socioeconomic deprivation.

AIM-HFpEF is designed to be fully automated and integrated into EHR platforms, has been externally validated and performs favourably to current diagnostic and screening models. It is anticipated that patients with a high likelihood of HFpEF as ascertained by the AIM-HFpEF model could be identified to the clinician by way of an electronic pop-up prompt within the EHR, with subsequent referral to a cardiologist for specialist assessment and initiation of treatment if appropriate.

In constructing AIM-HFpEF, we have taken the novel step of including data from patients with undiagnosed HFpEF in the datasets. We see this as a potential key approach in addressing the issue of underdiagnosis in HFpEF. Conceptually, it can be considered that the characteristics of the undiagnosed HFpEF patients are the clinically most important predictors, not captured through analysis solely of diagnosed HFpEF. One possible reason is that these patients not yet diagnosed have information missing in their structured data and only present in unstructured form. Patients within the Confirmed HFpEF group have *already been diagnosed* and therefore are in less of a need of a predictive model, whereas the patients within the Undiagnosed HFpEF group are those who are being missed by current diagnostic methods.

A key concern of AI-based disease prediction tools is the risk of potentiating any biases contained within the training dataset. A key aim of this study was to ensure that AIM-HFpEF retained good performance in diagnosing the significant minority of HFpEF patients of non-White race and ethnicity, to ensure alignment with future frameworks for assessing algorithmic bias ^20^. We note that the derivation and validation populations of the HFPEF-ABA score were overwhelming of White ethnicity and therefore its generalisability to non-White populations has not previously been ascertained. Furthermore, several studies have identified a lack of generalisability in the H2FPEF score *viz a viz* its performance in non-White populations, likely at least in part due to the heavy weighting afforded to a diagnosis of AF in the H2FPEF score (only a small proportion of Black patients with HFpEF have a diagnosis of AF) ^7^. Our results show that AIM-HFpEF performs better than H2FpEF and HFpEF-ABA in non-White patients in the UK, although we do note that both existing scores have reasonable performance in this patient group and our study can also be considered as additional external validation of these models.

The predictors identified in the full model can be related to HFpEF either in terms of direct pathophysiological mechanisms or by their relation to clinical features associated with the syndrome. In contrast to HFpEF-ABA, in our GLM model we found several echocardiogram variables to be significant predictors of HFpEF and therefore our model includes a number of echocardiogram measures i.e. Pulmonary artery systolic pressure (PASP), left atrial volume and LVEDD. The least explainable feature in our simplified model is TIA: we consider that it is likely to represent a composite of age, atrial fibrillation and cardiovascular disease, each of which are known to contribute to HFpEF pathophysiology. As expected, there is significant overlap between the variables included in AIM-HFpEF and those in other HFpEF predictive models. A key difference is the inclusion of NTproBNP in our model. Given that not all patients will have NTproBNP results available, we have confirmed the acceptable model performance even when natriuretic peptide results are not available (**Supplementary Document 1**).

### Strengths and Limitations

A key strength of an EHR-based approach is that it lends itself to automation i.e. there is no requirement for an *a priori* suspicion of HFpEF as is the case with diagnostic scores such as H2FPEF and HFA-PEFF. Conversely, the use of routinely collected, retrospective data does have key limitations, including non-standardised reporting of clinical features across the two participating centres, therefore requiring prospective validation.

As discussed above, the inclusion of patients with undiagnosed HFpEF in both the derivation and validation datasets is a strength of our approach, potentially allowing the final model to be much more generalisable and less prone to bias compared to diagnostic models derived from smaller, more highly preselected groups of confirmed HFpEF patients.

Another limitation of our model is the reliance on an advanced data extraction platform to employ NLP methods and retrieve clinical data from the EHR. Hospital systems with informatics capabilities to employ our model are in the minority globally, particularly in low- and lower-middle income countries, despite the CogStack technology being low cost and light weight, and available open source. We have sought to mitigate this limitation by producing an alternative prediction model that does not require advanced EHR data analytic capabilities. This **Simplified Model** could potentially be accessed via a smartphone app to enable clinicians to define the likelihood of a HFpEF diagnosis.

A further limitation is that although both the derivation and validation cohorts come from separate large multi-hospital NHS trusts, they are both within the same large urban metropolis i.e. London. Further external validation in different settings is therefore required to ensure generalisability of our findings across broader geographic areas. Further work will therefore involve assessment of wider generalisability both in larger UK datasets and in international datasets. Additional future avenues including prospective validation will be a key step toward assessing the ability of AIM-HFpEF to affect patient outcomes through improved diagnosis via the model. Finally, incorporation of primary care data will be important to ensure accurate diagnosis in the unknown proportion of undiagnosed HFpEF patients without clinical data within secondary care.

## Conclusion

In this study we describe the use of AI methods to develop an automated, EHR based diagnostic prediction model for HFpEF. The AIM-HFpEF model has been externally validated and is seen to perform favourably to existing diagnostic and screening models and is accurate in non-White patients and in those from areas of high socio-economic deprivation.

## Data availability statement

The data and code underlying this article will be shared on reasonable request to the corresponding author.

---

**Supplementary Figure 1.**
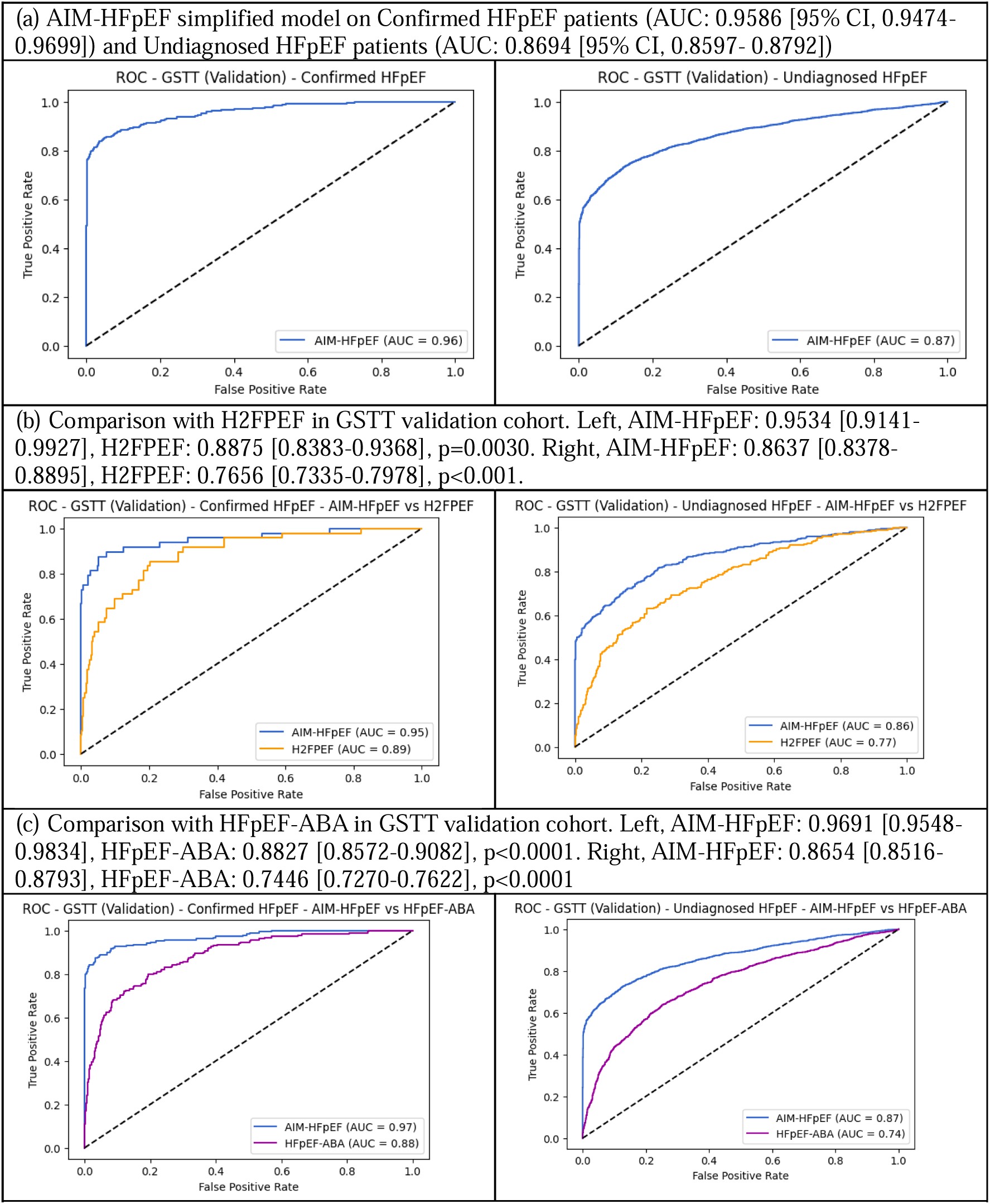
(a) Performance of the AIM-HFpEF prediction model on Confirmed HFpEF (left) and Undiagnosed HFpEF (right) patients in GSTT validation cohort. (b) Comparison with H2FPEF. (c) Comparison with HFpEF-ABA.

**Supplementary Figure 2.**
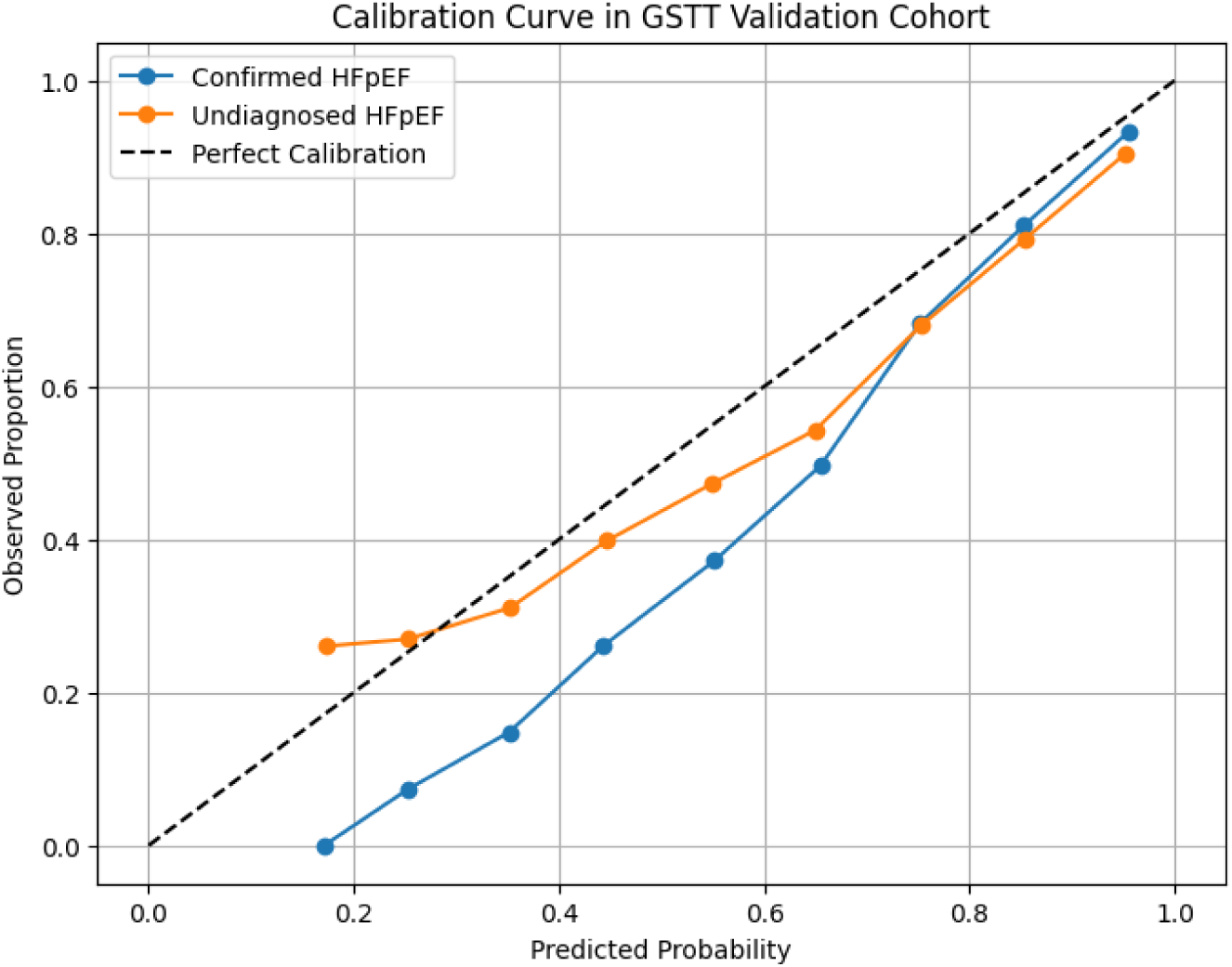
Calibration curves of predicted probabilities produced by the AIM-HFpEF simplified model. The undiagnosed HFpEF group is more calibrated, while the confirmed HFpEF group tends to have over-estimated probabilities when predicted probabilities are low.

**Supplementary Figure 3.**
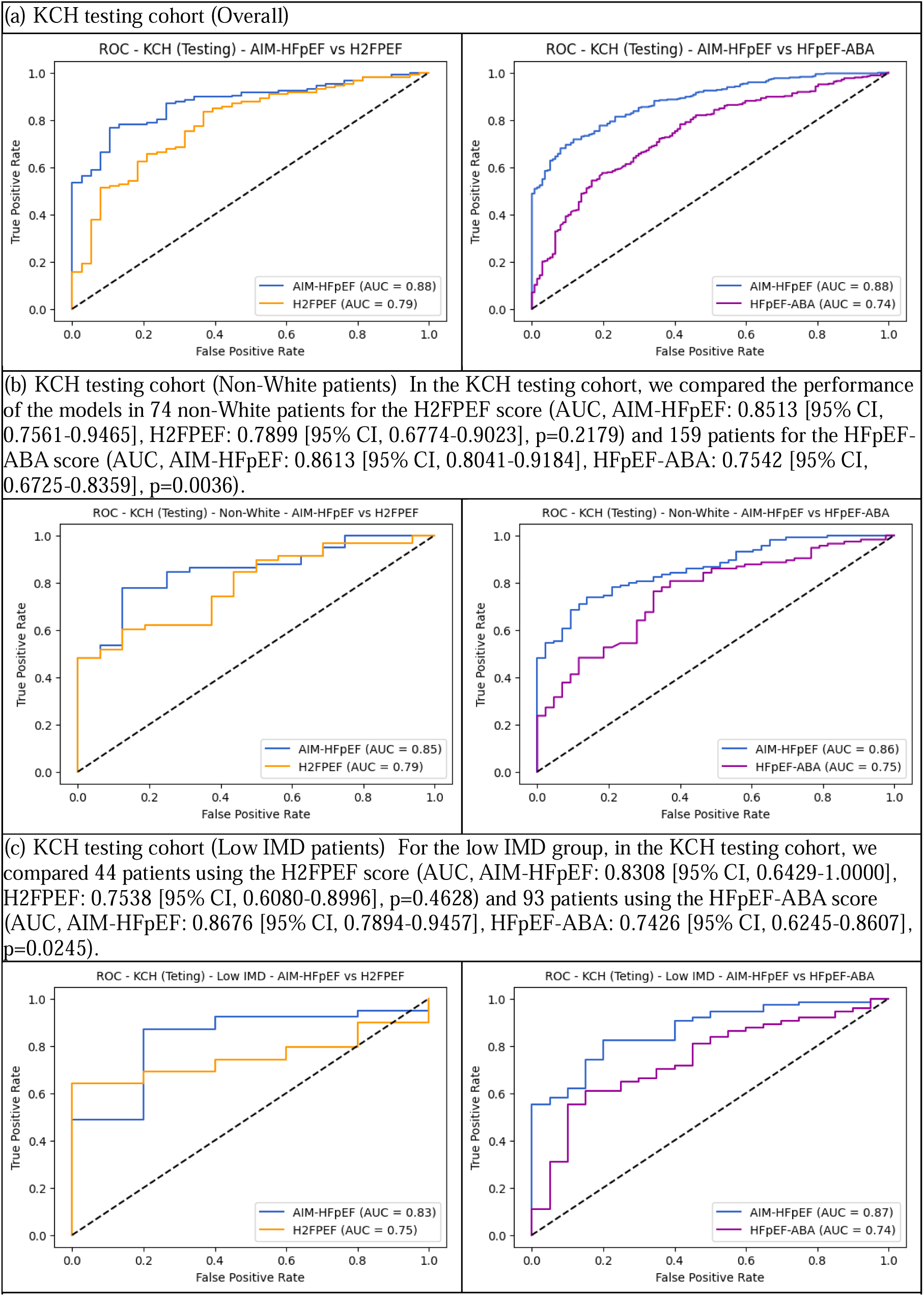
ROC curves for comparisons with H2FPEF and HFpEF-ABA scores in KCH testing cohort, (a) Overall (b) Non-White patients (c) Low IMD patients.

**Supplementary Figure 4.**
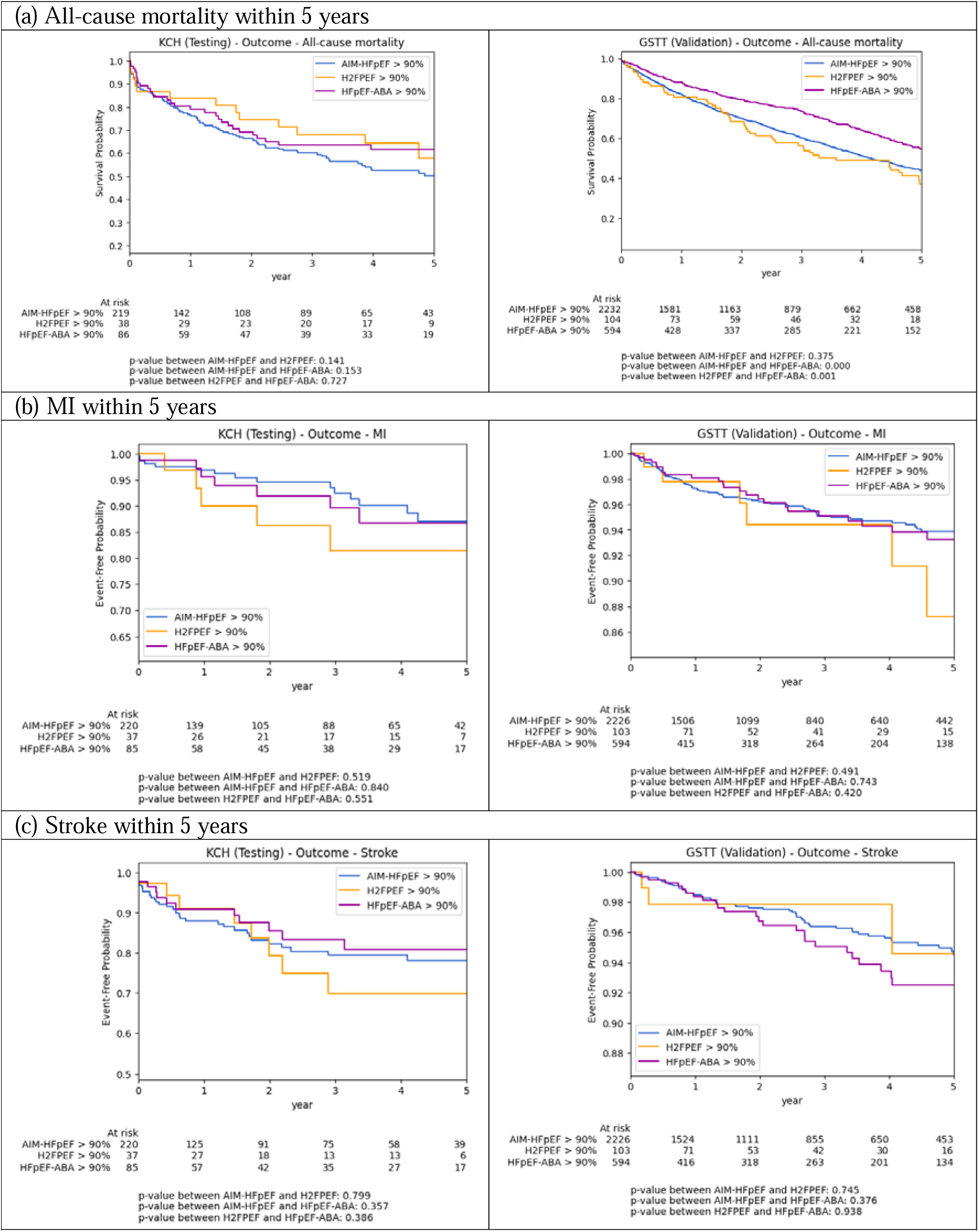
Kaplan-Meier (KM) curves for patients having a predicted probability ³ 90% of having HFpEF by the models with (a) all-cause mortality (b) MI and (c) Stroke within 5 years.

**Supplementary Table 1.** SNOMED-CT terms used in the study for the inclusion and exclusion criteria. All child terms are included by default. Inclusion criteria include HF, HFpEF and Dyspnoea. Exclusion criteria include severe valvular heart disease, hypertrophic cardiomyopathy, restrictive cardiomyopathy, constrictive pericarditis, and cardiac amyloidosis

| Criteria | SNOMED-CT Term | ID |
| --- | --- | --- |
| Inclusion | Heart failure (disorder) | 84114007 |
|  | Heart failure with normal ejection fraction (disorder) | 446221000 |
|  | Dyspnoea (finding) | 267036007 |
| Exclusion | Aortic valve stenosis (disorder)* | 60573004 |
|  | Aortic valve regurgitation (disorder)* | 60234000 |
|  | Mitral valve stenosis (disorder)* | 79619009 |
|  | Mitral valve regurgitation (disorder)* | 48724000 |
|  | Pulmonic valve stenosis (disorder)* | 56786000 |
|  | Pulmonic valve regurgitation (disorder)* | 91434003 |
|  | Hypertrophic cardiomyopathy (disorder) | 233873004 |
|  | Senile cardiac amyloidosis (disorder) | 16573007 |
|  | Restrictive cardiomyopathy (disorder) | 415295002 |
|  | Constrictive pericarditis (disorder) | 85598007 |

\* - the **severe** valvular heart disease patients are identified with the 'severe' keyword before the mentions of the SNOMED-CT terms.

**Supplementary Table 2.** Performances of all machine learning models.

| Model | Accuracy | Precision | Recall | F1-Score | AUC [95% CI] |
| --- | --- | --- | --- | --- | --- |
| F-LR | 0.7760 | 0.8094 | 0.8896 | 0.8476 | 0.8204 [0.7868-0.8540] |
| F-RF | 0.7382 | 0.7473 | 0.9459 | 0.8350 | 0.8472 [0.8161-0.8784] |
| F-SVM | 0.7918 | 0.8250 | 0.8919 | 0.8571 | 0.8386 [0.8063-0.8709] |
| F-XGB | **0.8186** | 0.8600 | 0.8851 | **0.8724** | **0.8910 [0.8665-0.9154]** |
| S-LR | 0.7539 | 0.7975 | 0.8694 | 0.8319 | 0.8025 [0.7678-0.8373] |
| S-RF | 0.7240 | 0.7307 | **0.9595** | 0.8296 | 0.8379 [0.8051-0.8707] |
| S-SVM | 0.7918 | 0.8223 | 0.8964 | 0.8578 | 0.8205 [0.7852-0.8559] |
| S-XGB | 0.8060 | **0.8607** | 0.8626 | 0.8616 | 0.8845 [0.8592-0.9097] |

KCH Testing Cohort – Comparison with H2FPEF (n=182)
| Model | Accuracy | Precision | Recall | F1-Score | AUC [95% CI] | P-value |
| --- | --- | --- | --- | --- | --- | --- |
| F-LR | 0.7935 | 0.8649 | 0.8767 | 0.8707 | 0.8398 [0.7764-0.9031] | 0.0753 |
| F-RF | 0.8043 | 0.8354 | 0.9384 | 0.8839 | 0.8600 [0.8060-0.9141] | 0.0318 |
| F-SVM | 0.7989 | 0.8658 | 0.8836 | 0.8746 | 0.8506 [0.7844-0.9168] | 0.0447 |
| F-XGB | 0.8261 | 0.8800 | 0.9041 | 0.8919 | **0.8756 [0.8245-0.9267]** | 0.0062 |
| S-LR | 0.7935 | 0.8699 | 0.8699 | 0.8699 | 0.8327 [0.7684-0.8971] | 0.0490 |
| S-RF | 0.8043 | 0.8274 | **0.9521** | 0.8854 | 0.8634 [0.8034-0.9233] | 0.0351 |
| S-SVM | 0.8152 | 0.8636 | 0.9110 | 0.8867 | 0.8131 [0.7350-0.8912] | 0.4362 |
| S-XGB | **0.8315** | 0.8912 | 0.8973 | **0.8942** | 0.8751 [0.8225-0.9276] | 0.0064 |
| H2FPEF | 0.6848 | **0.9231** | 0.6575 | 0.7680 | 0.7873 [0.7075-0.8672] | - |
| H2FPEF-Point | 0.8152 | 0.8544 | 0.9247 | 0.8882 | 0.7222 [0.6259-0.8184] | 0.1663 |

KCH Testing Cohort – Comparison with HFpEF-ABA (n=483)
| Model | Accuracy | Precision | Recall | F1-Score | AUC [95% CI] | P-value |
| --- | --- | --- | --- | --- | --- | --- |
| F-LR | 0.7734 | 0.8150 | 0.8837 | 0.8480 | 0.8206 [0.7821-0.8591] | <0.0001 |
| F-RF | 0.7443 | 0.7552 | 0.9506 | 0.8417 | 0.8454 [0.8096-0.8811] | <0.0001 |
| F-SVM | 0.7879 | 0.8288 | 0.8866 | 0.8567 | 0.8353 [0.7973-0.8732] | <0.0001 |
| F-XGB | **0.8170** | 0.8657 | 0.8808 | **0.8732** | **0.8873 [0.8587-0.9159]** | <0.0001 |
| S-LR | 0.7588 | 0.8081 | 0.8692 | 0.8375 | 0.7983 [0.7573-0.8392] | 0.0013 |
| S-RF | 0.7297 | 0.7388 | **0.9622** | 0.8359 | 0.8275 [0.7880-0.8671] | <0.0001 |
| S-SVM | 0.7963 | 0.8254 | 0.9070 | 0.8643 | 0.8148 [0.7724-0.8572] | 0.0007 |
| S-XGB | 0.8046 | **0.8676** | 0.8576 | 0.8626 | 0.8792 [0.8493-0.9091] | <0.0001 |
| HFpEF-ABA | 0.7173 | 0.8230 | 0.7703 | 0.7958 | 0.7425 [0.6949-0.7901] | - |

KCH Testing Cohort – Non-White (n=202)
| Model | Accuracy | Precision | Recall | F1-Score | AUC [95% CI] |
| --- | --- | --- | --- | --- | --- |
| F-LR | 0.7824 | 0.8108 | 0.8955 | 0.8511 | 0.8327 [0.7715-0.8938] |
| F-RF | 0.7668 | 0.7633 | **0.9627** | 0.8515 | 0.8576 [0.8044-0.9108] |
| F-SVM | 0.7927 | 0.8264 | 0.8881 | 0.8561 | 0.8443 [0.7852-0.9034] |
| F-XGB | **0.8135** | **0.8451** | 0.8955 | **0.8696** | **0.8848 [0.8390-0.9305]** |
| S-LR | 0.7513 | 0.7945 | 0.8657 | 0.8286 | 0.8034 [0.7397-0.8672] |
| S-RF | 0.7513 | 0.7529 | 0.9552 | 0.8421 | 0.8578 [0.8014-0.9143] |
| S-SVM | 0.7720 | 0.8041 | 0.8881 | 0.8440 | 0.8252 [0.7645-0.8859] |
| S-XGB | 0.7927 | 0.8406 | 0.8657 | 0.8529 | 0.8823 [0.8356-0.9290] |

KCH Testing Cohort – Non-White – Comparison with H2FPEF (n=74)
| Model | Accuracy | Precision | Recall | F1-Score | AUC [95% CI] | P-value |
| --- | --- | --- | --- | --- | --- | --- |
| F-LR | 0.7973 | 0.8909 | 0.8448 | 0.8673 | 0.8513 [0.7472-0.9554] | 0.1586 |
| F-RF | **0.8243** | 0.8571 | **0.9310** | **0.8926** | 0.8594 [0.7721-0.9467] | 0.1175 |
| F-SVM | 0.8108 | 0.9074 | 0.8448 | 0.8750 | 0.8524 [0.7477-0.9571] | 0.1786 |
| F-XGB | 0.8108 | 0.8793 | 0.8793 | 0.8793 | **0.8739 [0.7899-0.9580]** | 0.0614 |
| S-LR | 0.7703 | 0.8727 | 0.8276 | 0.8496 | 0.8222 [0.7160-0.9284] | 0.4025 |
| S-RF | 0.8108 | 0.8438 | **0.9310** | 0.8852 | 0.8815 [0.7954-0.9676] | 0.0559 |
| S-SVM | 0.7703 | 0.8475 | 0.8621 | 0.8547 | 0.8060 [0.6980-0.9141] | 0.7285 |
| S-XGB | 0.8108 | 0.8929 | 0.8621 | 0.8772 | 0.8513 [0.7561-0.9465] | 0.2179 |
| H2FPEF | 0.6622 | **0.9459** | 0.6034 | 0.7368 | 0.7899 [0.6774-0.9023] | - |
| H2FPEF-Point | 0.7703 | 0.8154 | 0.9138 | 0.8618 | 0.6546 [0.4886-0.8207] | 0.1400 |

KCH Testing Cohort – Non-White – Comparison with HFpEF-ABA (n=159)
| Model | Accuracy | Precision | Recall | F1-Score | AUC [95% CI] | P-value |
| --- | --- | --- | --- | --- | --- | --- |
| F-LR | 0.7707 | 0.8197 | 0.8772 | 0.8475 | 0.8027 [0.7268-0.8787] | 0.2133 |
| F-RF | 0.7707 | 0.7786 | **0.9561** | 0.8583 | 0.8355 [0.7721-0.8988] | 0.0420 |
| F-SVM | 0.7834 | 0.8390 | 0.8684 | 0.8534 | 0.8154 [0.7408-0.8900] | 0.1153 |
| F-XGB | **0.8089** | 0.8559 | 0.8860 | **0.8707** | **0.8694 [0.8142-0.9247]** | 0.0020 |
| S-LR | 0.7580 | 0.8167 | 0.8596 | 0.8376 | 0.7752 [0.6976-0.8528] | 0.5610 |
| S-RF | 0.7580 | 0.7676 | **0.9561** | 0.8516 | 0.8308 [0.7610-0.9006] | 0.0279 |
| S-SVM | 0.7707 | 0.8095 | 0.8947 | 0.8500 | 0.7997 [0.7260-0.8734] | 0.2027 |
| S-XGB | 0.7834 | 0.8448 | 0.8596 | 0.8522 | 0.8613 [0.8041-0.9184] | 0.0036 |
| HFpEF-ABA | 0.7197 | **0.8571** | 0.7368 | 0.7925 | 0.7542 [0.6725-0.8359] | - |

KCH Testing Cohort – Low IMD (n=118)
| Model | Accuracy | Precision | Recall | F1-Score | AUC [95% CI] |
| --- | --- | --- | --- | --- | --- |
| F-LR | 0.7812 | 0.8447 | 0.8788 | 0.8614 | 0.8213 [0.7416-0.9010] |
| F-RF | 0.8125 | 0.8319 | 0.9495 | 0.8868 | 0.8467 [0.7745-0.9190] |
| F-SVM | 0.8047 | 0.8700 | 0.8788 | 0.8744 | 0.8746 [0.8151-0.9341] |
| F-XGB | **0.8438** | 0.899 | 0.8990 | **0.8990** | **0.9011 [0.8460-0.9562]** |
| S-LR | 0.7656 | 0.8350 | 0.8687 | 0.8515 | 0.8001 [0.7177-0.8824] |
| S-RF | 0.7891 | 0.8000 | **0.9697** | 0.8767 | 0.8558 [0.7846-0.9270] |
| S-SVM | 0.7969 | 0.8763 | 0.8586 | 0.8673 | 0.8335 [0.7550-0.9120] |
| S-XGB | 0.8281 | **0.9053** | 0.8687 | 0.8866 | 0.8812 [0.8198-0.9427] |

KCH Testing Cohort – Low IMD – Comparison with H2FPEF (n=44)
| Model | Accuracy | Precision | Recall | F1-Score | AUC [95% CI] | P-value |
| --- | --- | --- | --- | --- | --- | --- |
| F-LR | 0.7727 | 0.9143 | 0.8205 | 0.8649 | 0.8359 [0.7065-0.9653] | 0.3105 |
| F-RF | 0.8409 | 0.8810 | 0.9487 | 0.9136 | 0.8077 [0.6738-0.9416] | 0.5139 |
| F-SVM | 0.7727 | 0.9143 | 0.8205 | 0.8649 | 0.8513 [0.7404-0.9622] | 0.1805 |
| F-XGB | 0.8409 | 0.9000 | 0.9231 | 0.9114 | 0.8615 [0.7282-0.9948] | 0.2082 |
| S-LR | 0.7955 | 0.9167 | 0.8462 | 0.8800 | 0.8513 [0.7320-0.9705] | 0.1345 |
| S-RF | 0.8636 | 0.8837 | **0.9744** | **0.9268** | **0.8923 [0.7955-0.9891]** | 0.0973 |
| S-SVM | 0.7955 | 0.9167 | 0.8462 | 0.8800 | 0.7795 [0.6015-0.9575] | 0.7758 |
| S-XGB | **0.8636** | 0.9459 | 0.8974 | 0.9211 | 0.8308 [0.6429-1.0000] | 0.4628 |
| H2FPEF | 0.6818 | **1.0000** | 0.6410 | 0.7812 | 0.7538 [0.6080-0.8996] | - |
| H2FPEF-Point | 0.7955 | 0.9167 | 0.8462 | 0.8800 | 0.6821 [0.3958-0.9683] | 0.5871 |

KCH Testing Cohort – Low IMD – Comparison with HFpEF-ABA (n=93)
| Model | Accuracy | Precision | Recall | F1-Score | AUC [95% CI] | P-value |
| --- | --- | --- | --- | --- | --- | --- |
| F-LR | 0.7660 | 0.8421 | 0.8649 | 0.8533 | 0.8034 [0.7023-0.9044] | 0.3102 |
| F-RF | 0.7979 | 0.8161 | **0.9595** | 0.8820 | 0.8007 [0.7034-0.8980] | 0.2763 |
| F-SVM | 0.7872 | 0.8553 | 0.8784 | 0.8667 | 0.8500 [0.7728-0.9272] | 0.0319 |
| F-XGB | **0.8404** | 0.8831 | 0.9189 | **0.9007** | 0.8892 [0.8185-0.9599] | 0.0098 |
| S-LR | 0.7660 | 0.8333 | 0.8784 | 0.8553 | 0.7682 [0.6585-0.8779] | 0.5925 |
| S-RF | 0.7979 | 0.8022 | 0.9865 | 0.8848 | 0.8324 [0.7381-0.9268] | 0.0784 |
| S-SVM | 0.7872 | 0.8462 | 0.8919 | 0.8684 | 0.8142 [0.7131-0.9152] | 0.1978 |
| S-XGB | 0.8298 | **0.8919** | 0.8919 | 0.8919 | **0.8676 [0.7894-0.9457]** | 0.0245 |
| HFpEF-ABA | 0.7234 | 0.8636 | 0.7703 | 0.8143 | 0.7426 [0.6245-0.8607] | - |

GSTT Validation Cohort (n=5,621)
| Model | Accuracy | Precision | Recall | F1-Score | AUC [95% CI] |
| --- | --- | --- | --- | --- | --- |
| F-LR | 0.7251 | 0.7364 | 0.8692 | 0.7973 | 0.7989 [0.7870-0.8108] |
| F-RF | 0.7485 | 0.7340 | 0.9343 | 0.8221 | 0.8348 [0.8237-0.8459] |
| F-SVM | 0.7377 | 0.7481 | 0.8719 | 0.8053 | 0.8086 [0.7970-0.8203] |
| F-XGB | **0.7986** | **0.8112** | 0.8815 | **0.8449** | **0.8934 [0.8852-0.9016]** |
| S-LR | 0.7269 | 0.7315 | 0.8865 | 0.8016 | 0.8053 [0.7936-0.8170] |
| S-RF | 0.7111 | 0.6998 | **0.9382** | 0.8016 | 0.8315 [0.8204-0.8426] |
| S-SVM | 0.7215 | 0.7288 | 0.8797 | 0.7972 | 0.7952 [0.7830-0.8073] |
| S-XGB | 0.7826 | 0.7986 | 0.8701 | 0.8328 | 0.8809 [0.8721-0.8897] |

GSTT Validation Cohort – Comparison with H2FPEF (n=967)
| Model | Accuracy | Precision | Recall | F1-Score | AUC [95% CI] | P-value |
| --- | --- | --- | --- | --- | --- | --- |
| F-LR | 0.6685 | 0.5817 | 0.8223 | 0.6814 | 0.7849 [0.7550-0.8149] | 0.6847 |
| F-RF | 0.6411 | 0.5493 | 0.9340 | 0.6917 | 0.8242 [0.7972-0.8513] | 0.0001 |
| F-SVM | 0.6783 | 0.5871 | 0.8553 | 0.6963 | 0.8016 [0.7726-0.8306] | 0.0623 |
| F-XGB | 0.7123 | 0.6105 | **0.9188** | **0.7335** | **0.8821 [0.8596-0.9047]** | <0.0001 |
| S-LR | 0.6783 | 0.5890 | 0.8401 | 0.6925 | 0.7815 [0.7514-0.8117] | 0.8918 |
| S-RF | 0.5722 | 0.5020 | 0.9569 | 0.6585 | 0.8071 [0.7788-0.8355] | 0.0199 |
| S-SVM | 0.6422 | 0.5548 | 0.8604 | 0.6746 | 0.7821 [0.7515-0.8128] | 0.8819 |
| S-XGB | 0.6860 | 0.5876 | 0.9112 | 0.7144 | 0.8746 [0.8510-0.8982] | <0.0001 |
| H2FPEF | **0.7144** | **0.6692** | 0.6675 | 0.6684 | 0.7805 [0.7505-0.8105] | - |
| H2FPEF-Point | 0.6772 | 0.5980 | 0.7665 | 0.6719 | 0.7550 [0.7242-0.7858] | 0.0072 |

GSTT Validation Cohort – Comparison with HFpEF-ABA (n=3,735)
| Model | Accuracy | Precision | Recall | F1-Score | AUC [95% CI] | P-value |
| --- | --- | --- | --- | --- | --- | --- |
| F-LR | 0.6391 | 0.5440 | 0.8496 | 0.6633 | 0.7800 [0.7644-0.7956] | 0.0119 |
| F-RF | 0.6446 | 0.5442 | 0.9255 | 0.6854 | 0.8236 [0.8098-0.8375] | <0.0001 |
| F-SVM | 0.6597 | 0.5610 | 0.8585 | 0.6786 | 0.7949 [0.7798-0.8101] | <0.0001 |
| F-XGB | **0.7518** | **0.6511** | 0.8763 | **0.7471** | **0.8898 [0.8784-0.9012]** | <0.0001 |
| S-LR | 0.6357 | 0.5399 | 0.8742 | 0.6675 | 0.7915 [0.7762-0.8068] | <0.0001 |
| S-RF | 0.5882 | 0.5042 | **0.9371** | 0.6557 | 0.8220 [0.8079-0.8362] | <0.0001 |
| S-SVM | 0.6340 | 0.5385 | 0.8742 | 0.6665 | 0.7886 [0.7731-0.8041] | <0.0001 |
| S-XGB | 0.7341 | 0.6331 | 0.8667 | 0.7317 | 0.8788 [0.8665-0.8912] | <0.0001 |
| HFpEF-ABA | 0.6623 | 0.5687 | 0.7977 | 0.6640 | 0.7624 [0.7462-0.7787] | - |

GSTT Validation Cohort – Non-White (n=1,182)
| Model | Accuracy | Precision | Recall | F1-Score | AUC [95% CI] |
| --- | --- | --- | --- | --- | --- |
| F-LR | 0.7125 | 0.7112 | 0.8610 | 0.7790 | 0.7956 [0.7703-0.8210] |
| F-RF | 0.7302 | 0.7019 | **0.9413** | 0.8042 | 0.8301 [0.8066-0.8536] |
| F-SVM | 0.7218 | 0.7201 | 0.8625 | 0.7849 | 0.8013 [0.7764-0.8261] |
| F-XGB | **0.7850** | **0.7814** | 0.8811 | **0.8283** | **0.8893 [0.8717-0.9070]** |
| S-LR | 0.7015 | 0.7014 | 0.8582 | 0.7719 | 0.7794 [0.7534-0.8053] |
| S-RF | 0.6855 | 0.6694 | 0.9198 | 0.7749 | 0.8166 [0.7926-0.8406] |
| S-SVM | 0.6813 | 0.6878 | 0.8395 | 0.7561 | 0.7616 [0.7343-0.7888] |
| S-XGB | 0.7572 | 0.7615 | 0.8553 | 0.8057 | 0.8641 [0.8437-0.8844] |

GSTT Validation Cohort – Non-White – Comparison with H2FPEF (n=249)
| Model | Accuracy | Precision | Recall | F1-Score | AUC [95% CI] | P-value |
| --- | --- | --- | --- | --- | --- | --- |
| F-LR | 0.7018 | 0.6439 | 0.8019 | 0.7143 | 0.7877 [0.7284-0.8469] | 0.4246 |
| F-RF | 0.6798 | 0.5976 | **0.9528** | 0.7345 | 0.8304 [0.7786-0.8823] | 0.0119 |
| F-SVM | 0.7149 | 0.6414 | 0.8774 | 0.7410 | 0.8017 [0.7447-0.8588] | 0.1444 |
| F-XGB | **0.7412** | 0.6556 | 0.9340 | **0.7704** | **0.9041 [0.8666-0.9416]** | <0.0001 |
| S-LR | 0.6886 | 0.6259 | 0.8208 | 0.7102 | 0.7674 [0.7062-0.8286] | 0.9653 |
| S-RF | 0.5965 | 0.5376 | 0.9434 | 0.6849 | 0.7856 [0.7265-0.8447] | 0.4863 |
| S-SVM | 0.6535 | 0.5931 | 0.8113 | 0.6853 | 0.7543 [0.6888-0.8199] | 0.5595 |
| S-XGB | 0.7018 | 0.6218 | 0.9151 | 0.7405 | 0.8812 [0.8365-0.9259] | <0.0001 |
| H2FPEF | 0.6974 | **0.7033** | 0.6038 | 0.6497 | 0.7681 [0.7071-0.8291] | - |
| H2FPEF-Point | 0.6711 | 0.6260 | 0.7264 | 0.6725 | 0.7394 [0.6760-0.8028] | 0.1688 |

GSTT Validation Cohort – Non-White – Comparison with HFpEF-ABA (n=923)
| Model | Accuracy | Precision | Recall | F1-Score | AUC [95% CI] | P-value |
| --- | --- | --- | --- | --- | --- | --- |
| F-LR | 0.6312 | 0.5326 | 0.8201 | 0.6458 | 0.7678 [0.7347-0.8010] | 0.0003 |
| F-RF | 0.6409 | 0.5350 | **0.9469** | 0.6837 | 0.8219 [0.7937-0.8500] | <0.0001 |
| F-SVM | 0.6590 | 0.5543 | 0.8584 | 0.6736 | 0.7898 [0.7581-0.8216] | <0.0001 |
| F-XGB | **0.7497** | **0.6387** | 0.8968 | **0.7460** | **0.8990 [0.8768-0.9212]** | <0.0001 |
| S-LR | 0.6215 | 0.5243 | 0.8289 | 0.6423 | 0.7569 [0.7231-0.7907] | <0.0001 |
| S-RF | 0.5816 | 0.4944 | 0.9145 | 0.6418 | 0.8031 [0.7725-0.8337] | <0.0001 |
| S-SVM | 0.6058 | 0.5119 | 0.8230 | 0.6312 | 0.7494 [0.7146-0.7841] | 0.0045 |
| S-XGB | 0.7170 | 0.6096 | 0.8614 | 0.7139 | 0.8744 [0.8477-0.9012] | <0.0001 |
| HFpEF-ABA | 0.6518 | 0.5609 | 0.6932 | 0.6201 | 0.7101 [0.6734-0.7468] | - |

GSTT Validation Cohort – Low IMD (n=1,136)
| Model | Accuracy | Precision | Recall | F1-Score | AUC [95% CI] |
| --- | --- | --- | --- | --- | --- |
| F-LR | 0.7324 | 0.7528 | 0.8670 | 0.8059 | 0.8074 [0.7816-0.8332] |
| F-RF | 0.7736 | 0.7580 | **0.9499** | 0.8432 | 0.8513 [0.8276-0.8751] |
| F-SVM | 0.7544 | 0.7731 | 0.8727 | 0.8199 | 0.8202 [0.7949-0.8454] |
| F-XGB | **0.8130** | **0.8322** | 0.8870 | **0.8587** | **0.8969 [0.8789-0.9148]** |
| S-LR | 0.7369 | 0.7470 | 0.8913 | 0.8128 | 0.8170 [0.7918-0.8422] |
| S-RF | 0.7287 | 0.7212 | 0.9399 | 0.8161 | 0.8376 [0.8137-0.8615] |
| S-SVM | 0.7324 | 0.7455 | 0.8841 | 0.8089 | 0.7993 [0.7724-0.8262] |
| S-XGB | 0.7837 | 0.8099 | 0.8655 | 0.8368 | 0.8811 [0.8615-0.9006] |

GSTT Validation Cohort – Low IMD – Comparison with H2FPEF (n=195)
| Model | Accuracy | Precision | Recall | F1-Score | AUC [95% CI] | P-value |
| --- | --- | --- | --- | --- | --- | --- |
| F-LR | 0.6902 | 0.6667 | 0.7609 | 0.7107 | 0.7811 [0.7133-0.8488] | 0.9449 |
| F-RF | 0.6957 | 0.6364 | 0.9130 | 0.7500 | 0.8355 [0.7751-0.8959] | 0.0365 |
| F-SVM | 0.7609 | 0.7264 | 0.8370 | 0.7778 | 0.8358 [0.7762-0.8954] | 0.0323 |
| F-XGB | **0.7935** | **0.7411** | 0.9022 | **0.8137** | **0.8982 [0.8505-0.9459]** | 0.0001 |
| S-LR | 0.7120 | 0.6726 | 0.8261 | 0.7415 | 0.7883 [0.7222-0.8544] | 0.6006 |
| S-RF | 0.6359 | 0.5817 | **0.9674** | 0.7265 | 0.8159 [0.7545-0.8773] | 0.1997 |
| S-SVM | 0.6793 | 0.6364 | 0.8370 | 0.7230 | 0.8029 [0.7366-0.8693] | 0.3887 |
| S-XGB | 0.7337 | 0.6720 | 0.9130 | 0.7742 | 0.8914 [0.8423-0.9406] | 0.0003 |
| H2FPEF | 0.7065 | 0.7317 | 0.6522 | 0.6897 | 0.7793 [0.7127-0.8459] | - |
| H2FPEF-Point | 0.7065 | 0.6759 | 0.7935 | 0.7300 | 0.7517 [0.6819-0.8214] | 0.1754 |

GSTT Validation Cohort – Low IMD – Comparison with HFpEF-ABA (n=726)
| Model | Accuracy | Precision | Recall | F1-Score | AUC [95% CI] | P-value |
| --- | --- | --- | --- | --- | --- | --- |
| F-LR | 0.6388 | 0.5488 | 0.8374 | 0.6630 | 0.7761 [0.7399-0.8123] | 0.9205 |
| F-RF | 0.6608 | 0.5602 | 0.9343 | 0.7004 | 0.8365 [0.8064-0.8665] | <0.0001 |
| F-SVM | 0.6799 | 0.5828 | 0.8651 | 0.6964 | 0.8058 [0.7721-0.8395] | 0.0459 |
| F-XGB | **0.7709** | **0.6736** | 0.8927 | **0.7679** | **0.8993 [0.8749-0.9237]** | <0.0001 |
| S-LR | 0.6344 | 0.5433 | 0.8685 | 0.6684 | 0.7949 [0.7599-0.8298] | 0.0832 |
| S-RF | 0.6050 | 0.5189 | **0.9481** | 0.6707 | 0.8240 [0.7927-0.8552] | 0.0013 |
| S-SVM | 0.6358 | 0.5443 | 0.8720 | 0.6702 | 0.7871 [0.7518-0.8224] | 0.3572 |
| S-XGB | 0.7342 | 0.6378 | 0.8651 | 0.7342 | 0.8829 [0.8557-0.9100] | <0.0001 |
| HFpEF-ABA | 0.6711 | 0.5799 | 0.8166 | 0.6782 | 0.7745 | - |

