## Supplementary Document 1 for "Artificial Intelligence Methods to Detect Heart Failure with Preserved Ejection Fraction (AIM-HFpEF) within Electronic Health Records: An equitable disease prediction model"

**Supplementary Document 1. AIM-HFpEF prediction model without NTproBNP**

This supplementary document presents the performance of the AIM-HFpEF prediction model, excluding the NTproBNP feature. This exclusion is the only modification compared to the full model in the main paper. **Figure 1** displays the SHAP graph for the model and the performance metrics for the KCH testing cohort (AUC, 0.8656 [95% CI, 0.8372-0.8940]) and the GSTT validation cohort (AUC, 0.8118 [95% CI, 0.8003-0.8234]).

| (a)  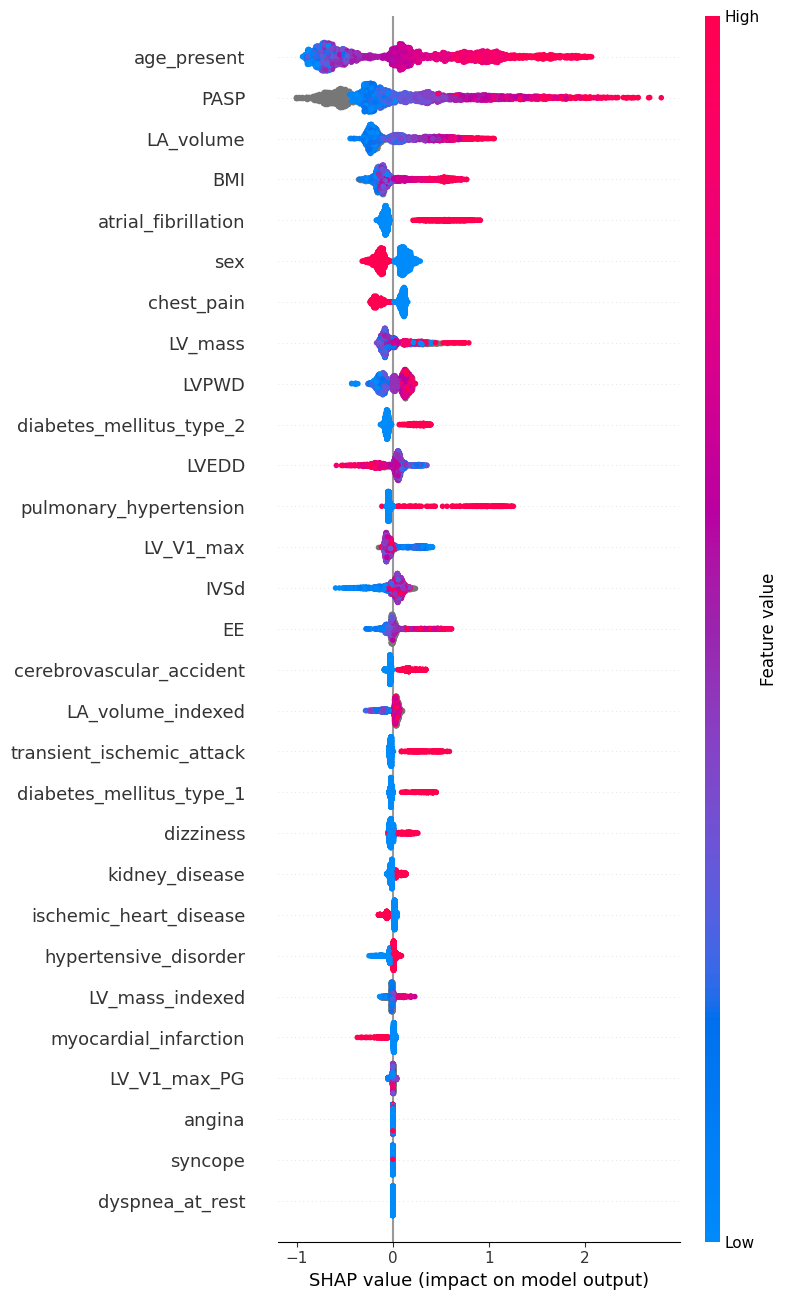 | (b)  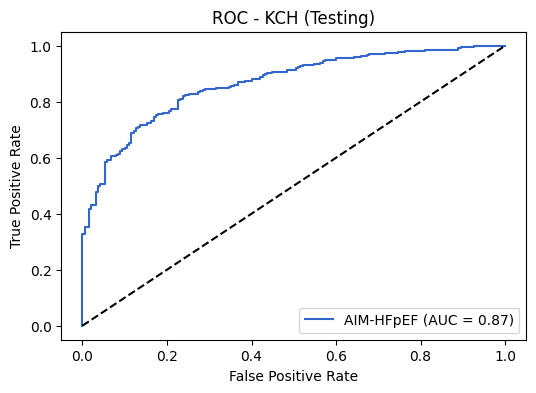 |
| --- | --- |
|  | (c)  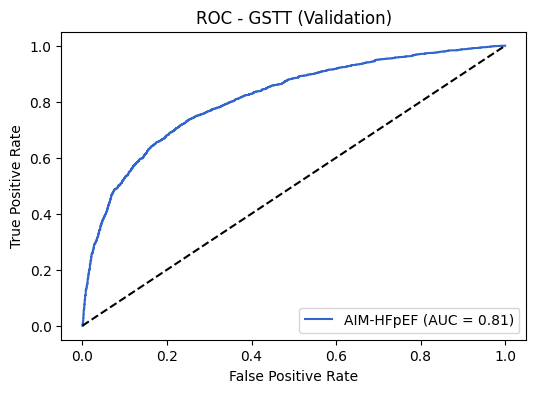 |
| Figure 1. (a) SHAP graph for AIM-HFpEF (full model without NTproBNP) (b) ROC curve of KCH testing cohort. (c) ROC curve of GSTT validation cohort. | |

*Comparison of Full Model with H2FPEF and HFpEF-ABA*

In the KCH testing cohort, the AUC of the Full Model for the 182 patients with the complete set of H2FPEF variables was 0.8398 [0.7775-0.9020], while the AUC of the H2FPEF score was 0.7873 [95% CI, 0.7075-0.8672], with a p-value of 0.0943. For the 483 patients with the full set of HFpEF-ABA variables, the AUC for AIM-HFpEF was 0.8610 [0.8278-0.8941], compared to the AUC for HFpEF-ABA, which was 0.7425 [95% CI, 0.6949-0.7901] (p<0.0001). **Figure 2 (a)** displays the corresponding ROC curves.

In the GSTT validation cohort, the AUC of the Full Model for the 483 patients with the complete set of H2FPEF variables was 0.8105 [95% CI, 0.7823-0.8387], while the AUC of the H2FPEF score was 0.7805 [95% CI, 0.7505-0.8105], with a statistically significant difference (p=0.0044). For the 3,735 patients with the full set of HFpEF-ABA variables, the AUC for AIM-HFpEF was 0.8055 [95% CI, 0.7909-0.8201], compared to the AUC for HFpEF-ABA, which was 0.7624 [95% CI, 0.7462-0.7787] (p<0.0001). **Figure 2 (b)** displays the corresponding ROC curves.

| (a) KCH Testing Cohort |  |
| --- | --- |
| 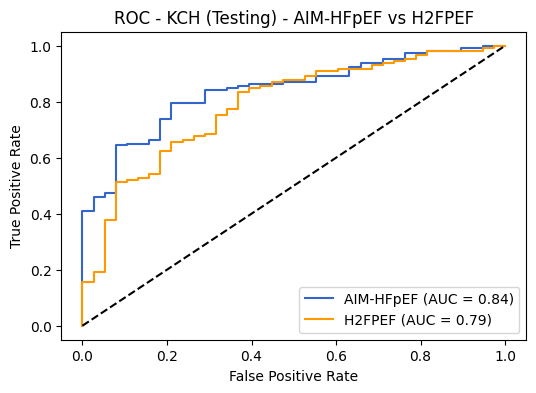 | 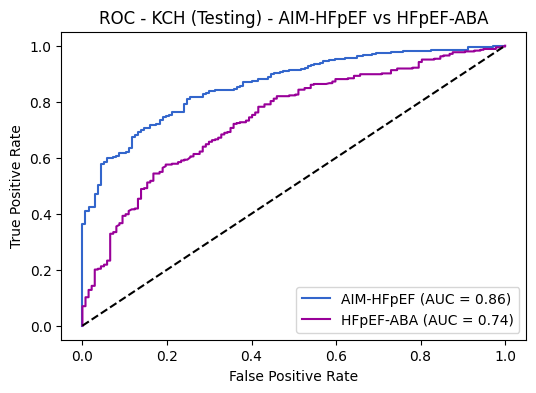 |
| (b) GSTT Validation Cohort |  |
| 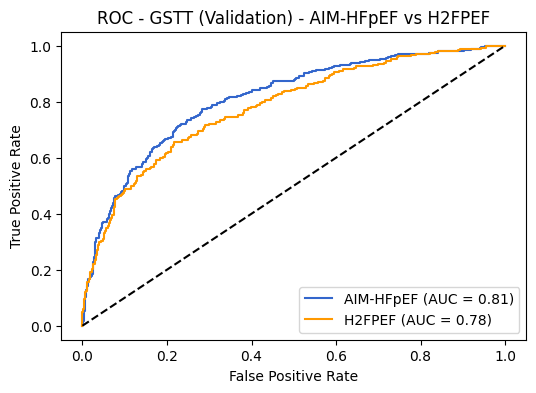 | 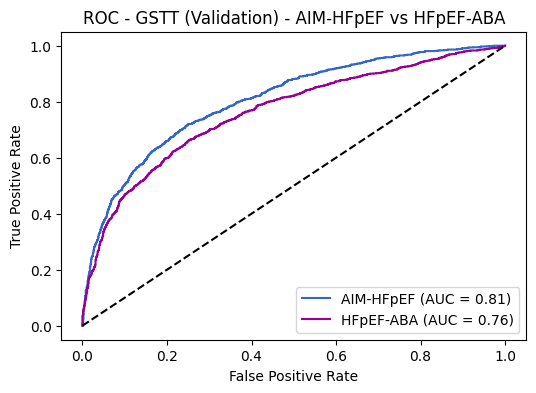 |
| Figure 2. Comparison of AIM-HFpEF (Full Model, without NTproBNP) with H2FPEF (left) and HFpEF-ABA (right) | |
